# A Time-Dependent Diffusion MRI Framework for Clinical Characterisation of Human Brain Cellular Architecture

**DOI:** 10.64898/2026.09.02.26362017

**Authors:** Anat Leibovici, Elena Espinós Soler, David Mesika, Galia Tsarfaty, Abigail Livny, Silvia De Santis, Maximilian F. Eggl

**Author notes:** co-corresponding authors (S.D. Santis); (M.F. Eggl).

## Abstract

Diffusion-weighted MRI, beyond the commonly used diffusion tensor framework, offers a unique window into tissue microstructure *in vivo*, yet its clinical adoption has remained limited. Major barriers include the complexity of diffusion MRI sequence design, lengthy acquisition protocols, and the challenges associated with robust estimation of high-dimensional microstructural model parameters. Here, we address these limitations by combining optimised diffusion encoding with state-of-the-art simulation-based inference, establishing a clinically feasible framework for multi-compartment diffusion modelling. We validate the approach through *i)* in-depth *in silico* experiments and *ii) in vivo* studies made up of both human and rodent data. The resulting microstructural metrics are robust, reproducible across healthy individuals and show significant spatial associations with brain-wide expression patterns of cell-specific genes. Requiring less than 10 minutes of acquisition time, this framework substantially lowers the barriers to advanced microstructural imaging, a prerequisite step toward its eventual evaluation for the diagnosis, stratification, and monitoring of brain disorders.

## 1. Introduction

Non-invasive measurement of cellular architecture in the living human brain remains a central challenge in neuroscience and clinical neurology, given that many neurological disorders are associated with microstructural abnormalities (Douaud et al., 2013; Zhan et al., 2012). Diffusion-weighted magnetic resonance imaging (dMRI) (Stejskal and Tanner, 1965; Le Bihan et al., 2001; Basser et al., 1994) gives access to brain microstructure *in vivo* and non-invasively, and has been applied in a variety of clinically relevant contexts, including acute brain ischaemia (Warach et al., 1992; Grand et al., 2013), infarct lesions (Chien et al., 1992), tumour detection (Koh and Collins, 2007), Alzheimer’s disease (Nir et al., 2024; Martínez-Tazo et al., 2026) and epilepsy (Liu et al., 2024).

While the standard diffusion tensor framework can be acquired with relatively modest imaging requirements, its pathological specificity is limited (De Santis et al., 2014). In contrast, denser sampling of the diffusion acquisition parameter space enables greater sensitivity and specificity to underlying pathological processes (Garcia-Hernandez et al., 2022; Palombo et al., 2020; Assaf and Basser, 2005; Assaf et al., 2008; Zhang et al., 2012; Jelescu et al., 2022; Olesen et al., 2022). Whereas some of these advanced microstructural models rely primarily on denser sampling of diffusion weightings and can therefore be readily integrated into clinical protocols (Fu et al., 2020; Schiavi et al., 2023), others require measurements with non-standard gradient pulses (Henriques et al., 2021; Yang et al., 2018; Westin et al., 2016), or across multiple diffusion times (Assaf et al., 2008; Jelescu et al., 2022; Olesen et al., 2022), substantially increasing acquisition complexity, thereby limiting clinical feasibility. In particular, probing diffusion-time dependence remains difficult on clinical scanners with moderate gradient strengths, where diffusion MRI is typically implemented using standard pulsed-gradient spin-echo sequences. As such, advanced microstructural characterisation remains largely confined to high-performance MRI systems (e.g., high-performance research platform or 7T scanners) (Foo et al., 2020; Jones et al., 2018) or preclinical scanners (Jelescu et al., 2020).

Even when a sequence can be implemented on a clinical MRI scanner, its practical utility in clinical studies is limited if the acquisition time exceeds even a few minutes. Clinical protocols typically require the inclusion of multiple standard imaging sequences and, increasingly, multimodal assessments. Consequently, prolonged acquisition times reduce feasibility, increase susceptibility to patient motion, and constrain adoption in routine clinical practice.

To address these challenges, we present a feasible clinical implementation of a dMRI protocol spanning both multiple diffusion weightings and times. We achieve this by combining optimised experimental design with modern fitting approaches based on simulation-based inference (SBI), which enables protocol optimisation and a substantial reduction in the number of required acquisitions (Eggl and De Santis, 2026).

Using a multi-compartment model inspired by the work of Garcia-Hernandez et al. (2022), designed to dissect glia contribution to dMRI signal, we first performed *in silico* experiments to define a clinically viable experimental protocol, taking into account scan duration and hardware constraints. Next, we demonstrated that the sequence implemented in a hospital setting 3T clinical scanner provides microstructural maps which retain most of the accuracy and precision as compared to the original implementation, while requiring less than 9 minutes to acquire. Finally, we show on healthy subjects that the cohort-averaged microstructural maps are significantly associated with microglia gene expression across brain regions. To demonstrate generalizability across modeling frameworks, we performed an exploratory analysis using the Neurite Exchange Imaging (NEXI) model to estimate exchange time (Jelescu et al., 2022). Taken together, our results bring advanced microstructural mapping within reach of routine clinical imaging in healthy individuals, motivating future studies to test its utility in the diagnosis and management of neurological diseases.

## 2. Methods

### 2.1. Multi-compartment model of diffusion

Here, we briefly describe the multi-compartment model employed in this study. A more comprehensive treatment is provided by Garcia-Hernandez et al. (2022). In this model, the underlying substrate is treated as a set of discrete compartments, each exhibiting different diffusion behaviors. By defining compartments that approximate, for example, the structure of neurons or glial cells, we aim to obtain biomarkers that provide crucial insight not available via the simple mathematical treatments of the diffusion signal. In the model of this work, we defined three distinct microstructural compartments: a hindered compartment (denoted by (⋅)_*h*_), a spherical compartment ((⋅)_*s*_), and a cylindrical compartment ((⋅)_*c*_), as illustrated in Fig. S1a. In contrast to the original of Garcia-Hernandez et al. (2022), who used two separate spherical compartments to model microglia and astrocytes, we introduced this simpler single spherical compartment due to clinical considerations, including the need to reduce the number of diffusion-weighting shells required for model fitting. Additionally, as spheres and cylinders are not all of one uniform size, we modelled each of these distributions of sizes using a Poisson distribution, similar to previous work (De Santis et al., 2016b; Eggl and De Santis, 2026).

The normalised diffusion signal is therefore expressed as

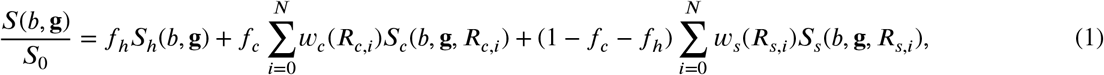

where *S*(*b*, **g**) denotes the diffusion-weighted signal acquired with diffusion weighting *b* and gradient direction **g**, and *S*_0_ is the corresponding non-diffusion-weighted signal. The parameters *f*_*h*_ and *f*_*c*_ denote the volume fractions of the hindered and cylindrical compartments, respectively, with the spherical compartment fraction given by 1 − *f*_*c*_ − *f*_*h*_. The terms *S*_*h*_, *S*_*c*_, and *S*_*s*_ represent the corresponding compartment signals, whereas *w*_*c*_ and *w*_*s*_ denote the normalised Poisson weights associated with the cylindrical and spherical radius distributions, respectively. Finally, *N* defines how finely we want to subdivide our Poisson distribution - in this work *N* = 100.

#### 2.1.1. Models with exchanging compartments

As we also investigate the suitability of probing the diffusion between compartments, we briefly describe the NEXI model introduced by Jelescu et al. (2022). NEXI is a two-compartment model comprising an extracellular compartment with isotropic diffusion and a stick-like intracellular compartment, in which diffusion is restricted to the direction parallel to the neurites, which are assumed to be infinitely thin. Critically, rather than treating these compartments as isolated diffusion environments, NEXI assumes exchange of water across permeable compartment boundaries. Measurements at different diffusion times can therefore provide sensitivity to the characteristic exchange time, *t*_ex_.

Under the short-gradient-pulse approximation, NEXI reduces to an analytical form that enables straightforward simulation and fitting of *t*_ex_.

### 2.2. Simulation-based inference implementation

In this work, SBI was used to implement the model in Eq. 1 to achieve two objectives: *i)* to determine a clinically compatible experimental protocol using *in silico* validation and *ii)* use this protocol to extract microstructural maps from *in vivo* human and rodent data.

We employed neural posterior estimation (NPE) (Lueckmann et al., 2017, 2021), an SBI method, to estimate the parameters of the stick-and-ball model. NPE uses normalising flows (Trippe and Turner, 2018), a class of flexible probabilistic models obtained by applying a sequence of invertible transformations to simple base distributions such as Gaussians. By parameterising these transformations with neural networks, NPE can learn an accurate approximation of the posterior distribution over model parameters, even when the likelihood function is intractable.

Fig. S1d provides a schematic overview of the NPE procedure. During training, candidate parameters are sampled from a prior distribution and used to generate synthetic signals with the chosen forward models. The NPE network is trained to approximate the posterior distribution over model parameters conditioned on the simulated signals by minimising the negative log probability of the sampled parameters under the predicted posterior. After training, experimental data is passed through the network to obtain the posterior distribution of the model parameters. Because the network outputs a full posterior distribution, it naturally provides a measure of parameter uncertainty conditioned on both the observed data and the specified prior. Several posterior summaries can be used to obtain point estimates, including the posterior mean, posterior mode, or maximum a posteriori (MAP) estimate (Bassett and Deride, 2019). In this work, we used the posterior mean, computed efficiently by averaging a large batch of posterior samples. This approach avoids additional voxel-wise optimisation while providing a stable estimate of the posterior central tendency.

Based on biophysically plausible values reported in the literature, prior ranges were defined for all model parameters (e.g. for the stick diameters (Huang et al., 2019)). Synthetic diffusion signals were then generated by evaluating the forward multi-compartment model using parameters sampled from these ranges. A total of 250,000 simulations were generated with Rician noise corresponding to an SNR of 20 and used to train the NPE network - in line with results found in Eggl and De Santis (2026). All simulation-based inference analyses were performed using the SBI toolbox (version 0.24.0) (Tejero-Cantero et al., 2020). This analysis and accompanying Jupyter notebooks can be found in the following GitHub repository.

### 2.3. Implementation of the multi–diffusion-time framework to a clinically feasible human protocol

The multi-diffusion-time (multi-Δ) stick-and-ball framework was originally developed for high-gradient preclinical imaging in rodents and tested on a human 3T system with enhanced gradient capabilities (Garcia-Hernandez et al., 2022). To enable in vivo human application on a clinical 3T system, the acquisition protocol was adapted to satisfy gradient hardware limitations, safety constraints, and clinically feasible scan times while preserving the underlying modelling framework.

Three diffusion times (Δ = 27, 40, and 60 ms) were selected to probe distinct diffusion length scales while maintaining acceptable echo times (TEs) and gradient duty-cycle constraints. Although shorter diffusion times were used in the original preclinical implementation (Garcia-Hernandez et al., 2022), the minimum achievable diffusion time on the clinical 3T system was limited by gradient hardware constraints.

Diffusion timing parameters (Δ and δ) were adjusted using the Siemens work-in-progress (WIP) Advanced Diffusion sequence. Important parameters are reported in Table 1.

**Table 1:** Acquisition parameters for the multidiffusion-time protocol implemented on a hospital setting 3T clinical scanner.

| Parameter | Value |
| --- | --- |
| $\delta$ | 22 ms |
| TE | 101 ms |
| TR | 13.6 s |
| Number of slices | 74 |
| Voxel size | 2 mm<sup>3</sup> |
| FOV | 220 |
| bval | 2000 / 4000 |
| directions per shell | 6 |
| $b_0$ | 1 image |
| 13 acquisitions per $\Delta$ | |
| 39 acquisitions in 9 minutes |  |

To maximize diffusion weighting within the gradient amplitude limits of the hospital setting clinical platform system, diffusion encoding was performed using two orthogonal gradient axes, allowing each axis to operate at its maximum permitted amplitude. This configuration yields an effective gradient magnitude of 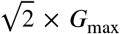, thereby increasing diffusion weighting without exceeding per-axis hardware constraints (Fig. S1b). The resulting geometric constraints on the diffusion encoding vectors limited the acquisition to six unique diffusion directions per shell (Fig. S1c). The sequence was implemented within vendor-approved safety limits, with automatic recalculation of the b-matrix.

### 2.4. Participants

This study was conducted in accordance with the Declaration of Helsinki and received ethical approval from the Ministry of Health (approval number 20186947) and the Sheba Medical Center Ethics Committee (approval number SMC-16-3035). Written informed consent was obtained from all participants prior to study participation. All structural MRI scans were independently reviewed by a board-certified neuroradiologist, and no clinically significant abnormalities were identified.

Participants had no history of neurological or psychiatric disorders. No significant age differences were observed between sexes (one-way ANOVA, *F* (1, 22) = 0.002, *p* = 0.962; Table S1). Of the 24 enrolled participants, one was excluded prior to analysis because mean head motion exceeded the pre-specified threshold of 0.4 mm (0.443 mm), resulting in a final sample of 23 participants. Among the included participants, mean volume-to-volume motion was 0.129 ± 0.082 mm (range, 0.028–0.328 mm), and maximum motion was 0.805 ± 0.751 mm (range, 0.156–2.758 mm). Full motion metrics for all participants are provided in Supplementary Table S2.

No participants met the predefined intensity outlier criterion on any metric. Across the 23 included participants, *z* |*z*| >. all spatial mean and spatial standard deviation-scores remained below the exclusion threshold of 2 5. The maximum absolute *z*-score observed across all metrics was 2.20, indicating stable parameter estimation and the absence of gross reconstruction failures (Supplementary Table S3). Visual inspection further confirmed the absence of signal dropout, echo-planar imaging (EPI) distortion artefacts, and gross co-registration errors in all included participants.

### 2.5. MRI Data Acquisition

All participants underwent MRI scanning on a 3T Siemens MAGNETOM Prisma scanner at the Division of Diagnostic Imaging, Sheba Medical Center. Structural imaging included a high-resolution T1-weighted magnetization-prepared rapid gradient-echo (MP-RAGE) sequence (TR = 2100 ms, TE = 2.45 ms, TI = 900 ms, flip angle = 8°, field of view = 256 mm, isotropic voxel size = 1 mm^3^). Fluid-attenuated inversion recovery (FLAIR) and susceptibility-weighted imaging (SWI) sequences were additionally acquired for clinical evaluation and exclusion of incidental pathology.

Conventional diffusion-weighted imaging was performed using a spin-echo EPI sequence for derivation of fractional anisotropy (FA) and mean diffusivity (MD). Acquisition parameters were 72 axial slices, TR = 2400 ms, TE = 66.4 ms, flip angle = 78°, field of view = 172 × 224 mm^2^, and isotropic voxel size = 2 mm^3^. A multi-shell diffusion protocol was acquired with *b* = 350, 1000, and 2500 s/mm^2^ using 32, 64, and 64 diffusion-encoding directions, respectively, together with four non-diffusion-weighted (*b* = 0) images. Multi-band EPI acquisition (acceleration factor = 3) was implemented using sequences developed by the Center for Magnetic Resonance Research.

The multi-Δ sequence was applied using the same geometry of the conventional diffusion-weighted sequence, but the experimental scheme described in section 2.3. The acquisition time for this sequence was 9 minutes.

Diffusion MRI preprocessing was performed semi-automatically using MRtrix3 (Tournier et al., 2019), MATLAB (R2022b), the FMRIB Software Library (FSL, v6.0.1), and custom Bash scripts. The preprocessing pipeline comprised three stages. First, diffusion signal denoising was performed using a random matrix theory–based approach (Veraart et al., 2016). Second, correction for subject motion, eddy-current-induced distortions, and susceptibility-induced geometric distortions was performed using FSL `topup` and `eddy` (Andersson et al., 2003; Andersson and Sotiropoulos, 2016), implemented through the MRtrix3 wrapper `dwifslpreproc` (Tournier et al., 2019), with phase-encoding direction `j-` and the `eddy` option `–slm=linear`. Eddy quality-control outputs were generated for all subjects using the `-eddyqc_all` option.

To ensure consistent motion correction across diffusion times for the multi-Δ sequence, volumes acquired at all three diffusion times were concatenated into a single 4D dataset comprising 39 volumes prior to preprocessing. Because all diffusion-time blocks were acquired sequentially within a single scanning session without table repositioning, motion and distortion correction were applied jointly to the concatenated dataset, thereby correcting both intra- and inter-block motion within a common reference frame. Bias-field correction was subsequently performed to account for B1 field inhomogeneity.

MD and FA maps were extracted from the multi-shell acquisition using an in-house Python code (version 3.8). The SBI toolbox (version 0.24.0) (Tejero-Cantero et al., 2020) Python package was used as a basis for an in-house implementation to extract microstructural maps from the multi-Δ acquisition.

#### 2.6. Statistical Analysis

Statistical analyses of the *in silico* experiments, the external validation dataset (Garcia-Hernandez et al., 2022), and the *in vivo* experiments were performed to evaluate the accuracy, precision, and robustness of parameter estimates obtained using the SBI framework.

#### 2.6.1. In silico validation

For *in silico* experiments, each replicate corresponded to an independently simulated diffusion signal generated from a randomly sampled parameter set and an independent noise realisation. Unless otherwise stated, quantitative results were computed from 1,000 simulated test signals. Performance was assessed by comparing reconstructed diffusion signals and estimated model parameters with their corresponding ground-truth values.

#### 2.6.2. Validation on high-performance MRI dataset

The external human diffusion MRI dataset comprised 29 acquisitions from six participants, each scanned on a high-gradient high-performance research platform scanner with an additional 2–4 test–retest sessions, as reported by Garcia-Hernandez et al. (2022). Each acquisition session was treated as an independent replicate.

Because no ground truth is available for *in vivo* human data, a reduced acquisition protocol, designed to approximate the clinically feasible protocol used in this study, was compared with its respective full acquisition protocol. The reduced protocol consisted of three diffusion times (Δ = 17, 35, and 61 ms) with six diffusion directions per b-shell (b = 2000, b = 4000 s/mm^2^) chosen to maximize electrostatic repulsion, whereas the full protocol comprised 273 acquisitions (1 *b*_0_ image, 30 directions at *b* = 2000s/mm^2^, and 60 directions at *b* = 4000 s/mm^2^ across all diffusiontime conditions). Agreement between the reduced- and full-protocol estimates was evaluated on a voxel-wise basis using normalised bias, root-mean-square error (RMSE), and Pearson correlation.

For test–retest analyses, repeated sessions from the same participant were treated as within-subject replicates. Reproducibility was quantified using the within-subject coefficient of variation (wCV) for selected model parameters. Summary statistics are reported as medians and interquartile ranges unless otherwise stated.

Additionally, region-wise analysis was performed at the level of six macro-regions, with hemispheres bilaterally aggregated. These six macro-regions were defined as cerebral cortex, cerebellum, thalamus, basal ganglia, hippocampal formation and amygdala.

The rat data from the same dataset were used to assess sensitivity of the model to inflammation, following the approach of Garcia-Hernandez et al. (2022). Lesions induced by lipopolysaccharide (LPS) injection were compared with contralateral regions injected with the same amount of saline within the same animal using two-sided Mann–Whitney U tests. Multiple comparisons were controlled using a family-wise error rate of α = 0.05. Asymmetry indices were computed for each animal and summarised across the cohort.

#### 2.6.3. Analysis of novel clinical dataset

Similarly to the high-gradient dataset, no ground truth was available for the clinical *in vivo* human data. Therefore, an extended acquisition protocol was used as a reference standard for evaluating the clinically feasible protocol. To accommodate the geometric constraints of the gradient-boosting implementation, 12 signed diffusion directions were acquired per b-shell and repeated once, yielding 24 acquisitions per *b*-shell and a total of 144 acquisitions acquired over approximately 30 min (*n* = 6). Parameter estimates obtained from this extended protocol were compared with the clinically feasible six-direction protocol through subsampling the same participant using the metrics described above. Unless otherwise stated, inferential statistical analyses were performed at the level of eight predefined macroregions, with bilateral hemispheres aggregated. The macro-regions comprised the cerebral cortex, cerebellum, thalamus, basal ganglia, hippocampal formation, amygdala, midbrain, and pons–medulla. Details of the brain parcellation are provided in the Supplementary Methods.

Multiple linear regression was used to assess associations between study variables, with imaging-derived predictors entered jointly in each model. Model performance was summarised using the overall *F* statistic, *R*^2^, and adjusted *R*^2^. For each predictor, standardised regression coefficients (β), standard errors, 95% confidence intervals, and partial *R*^2^ values were reported. Agreement between observed and predicted transcriptional values was quantified using Pearson correlation. To account for the limited number of macro-regions, non-parametric bootstrap resampling (10,000 iterations) was performed to derive 95% confidence intervals and empirical *p*-values. Sex differences in age were assessed using one-way ANOVA. All statistical analyses were conducted in Python (version 3.8).

Human microglial transcriptional data were obtained from the publicly available Human Brain Cell Atlas (HBCA) non-neuronal dataset (Human Cell Atlas Data Portal, accessed November 30, 2025) and organised according to the transcriptomic taxonomy described by Siletti et al. (2023). Analyses were restricted to cells annotated as microglia and summarised using the anatomical region labels provided in the HBCA metadata. MRI-derived macro-regions were subsequently harmonized with the corresponding HBCA anatomical definitions to enable cross-modal comparison between imaging and transcriptional measurements. To reduce sampling bias arising from unequal cell counts across donors and regions, transcriptional values were first averaged within each donor and region and subsequently averaged across donors to obtain region-level estimates.

## 3. Results

### 3.1. In silico validation

To assess whether the SBI framework can compensate for the limitations imposed by reduced acquisition time (necessary for clinical implementation) and inhomogeneous angular sampling (imposed by the reduced number of unique diffusion directions per shell available), we evaluated performance across varying levels of acquisition subsampling using simulated data with known ground-truth parameters. As shown in Fig. 1a,b, acceptable signal reconstruction performance was maintained with as few as six acquisitions per shell, both for a representative example signal and across 1000 simulated test cases quantified using RMSE and Pearson correlation. This finding is consistent with previous observations reported by Eggl and De Santis (2026) using the AxCaliber framework.

**Figure 1:**
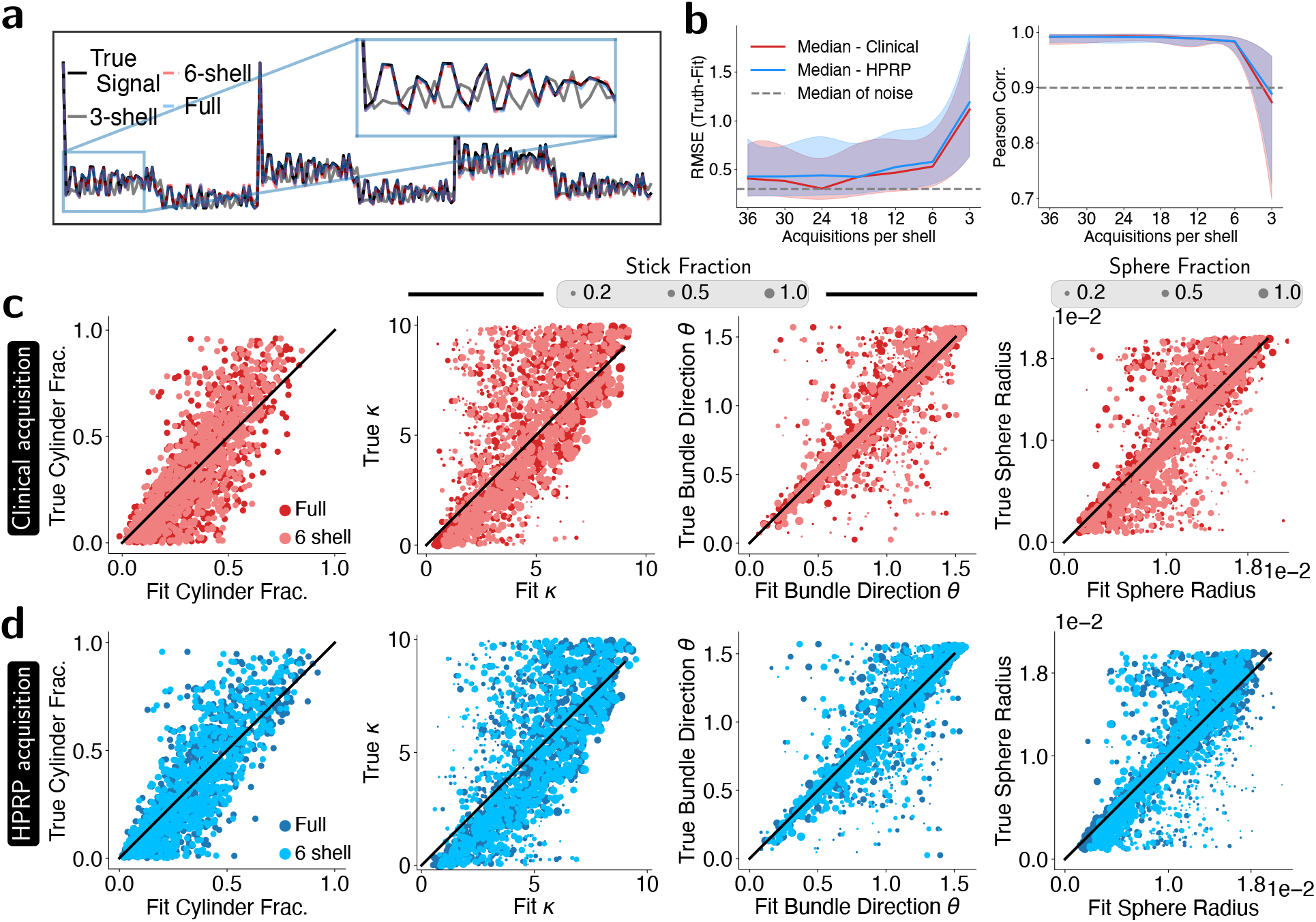
Simulation-based inference enables reliable estimation of multi-Δ microstructural parameters under clinically feasible acquisition conditions. **a)** For a randomly selected parameter set, a ground-truth diffusion signal (black) was generated. A noisy realisation was then provided to the network, and reconstruction performance was evaluated under different acquisition densities (full acquisition, blue; six acquisitions per shell, red; three acquisitions per shell, gray). **b)** Reconstruction accuracy across acquisition schemes, quantified using RMSE (left) and Pearson correlation (right) relative to the ground-truth signal, for the hospital setting clinical platform (minimum Δ = 27 ms, red) and high-performance research platform (minimum Δ = 17 ms, blue; (Garcia-Hernandez et al., 2022)) protocols. The median RMSE between the noisy and ground-truth signals is shown for reference. A Pearson correlation of 0.9 is shown for reference. Solid lines denote medians and shaded regions indicate the interquartile range (IQR). **c**,**d)** Comparison of true and inferred parameters for the full acquisition (dark colors) and reduced acquisition (six acquisitions per shell, lighter colors) using the hospital setting clinical platform (c, red) and high-performance research platform (d, blue) protocols. For the orientation-dispersion parameter κ, marker size is proportional to the cylinder fraction. The black line denotes the identity line (*y* = *x*).

We next evaluated parameter recovery across a representative range of randomly sampled parameter configurations (Fig. 1c) and compared the proposed clinical acquisition scheme with the high-performance research platform-based protocol of Garcia-Hernandez et al. (2022), which included shorter diffusion times inaccessible under standard gradient regime (Fig. 1d). Motivated by their findings, which identified cylinder fraction *f*_*c*_, orientation dispersion κ, and sphere radius λ_*s*_ as particularly sensitive markers of microglial reactivity, we focused our analysis on these parameters.

Overall, the two protocols exhibited comparable performance across model parameters for both the full and six-direction acquisition schemes. The lowest estimation accuracy of the hospital setting clinical platform was observed for κ (Pearson correlation = 0.59). Most deviation errors occurred when the *f*_*c*_ was below 0.2, a regime in which the cylindrical compartment contributes only minimally to the measured diffusion signal and reliable estimation of orientation dispersion becomes challenging. Restricting the analysis to voxels with *f*_*c*_ greater than 0.2 improved the correlation for κ to 0.77. As expected, the principal difference between the hospital setting clinical and highperformance research platform protocols was observed for λ_*s*_ estimation, where the latter acquisition showed improved performance for the smallest sphere radii owing to its access to shorter diffusion times.

These results demonstrate that the limitations imposed by clinical hardware, including restricted angular sampling and longer minimum diffusion times, can be largely mitigated through SBI-based parameter estimation. Together, these findings support the feasibility of translating the preclinically validated multi-Δ framework to routine human imaging on a hospital setting clinical platform.

### 3.2. In vivo validation

We next evaluated whether the performance observed in the *in silico* experiments generalized to *in vivo* data using the high-gradient high-performance research platform dataset of Garcia-Hernandez et al. (2022). As described in the methods, the original dataset was retrospectively downsampled to six diffusion directions per *b*-shell.

Representative parameter maps from a medial axial slice of a single subject, reconstructed using either the full acquisition protocol (273 acquisitions across three diffusion times) or the reduced protocol (six acquisitions per *b*-shell), are shown in Fig. 2a–c. Despite the substantial reduction in sampling density (86% fewer acquisitions), the reduced protocol preserved the major anatomical features observed in the full-acquisition maps, albeit with some loss of fine structural detail. We next repeated this analysis in a participant from the hospital setting clinical dataset for whom an extended 24-acquisition-per-shell protocol was available. Consistent with the results obtained in the high-performance research platform dataset, the six-direction protocol preserved the major anatomical features observed in the extended acquisition, despite the substantial reduction in sampling density. The principal differences were limited to a loss of fine structural detail and increased local variability in the parameter maps (Fig. 2d–f).

**Figure 2:**
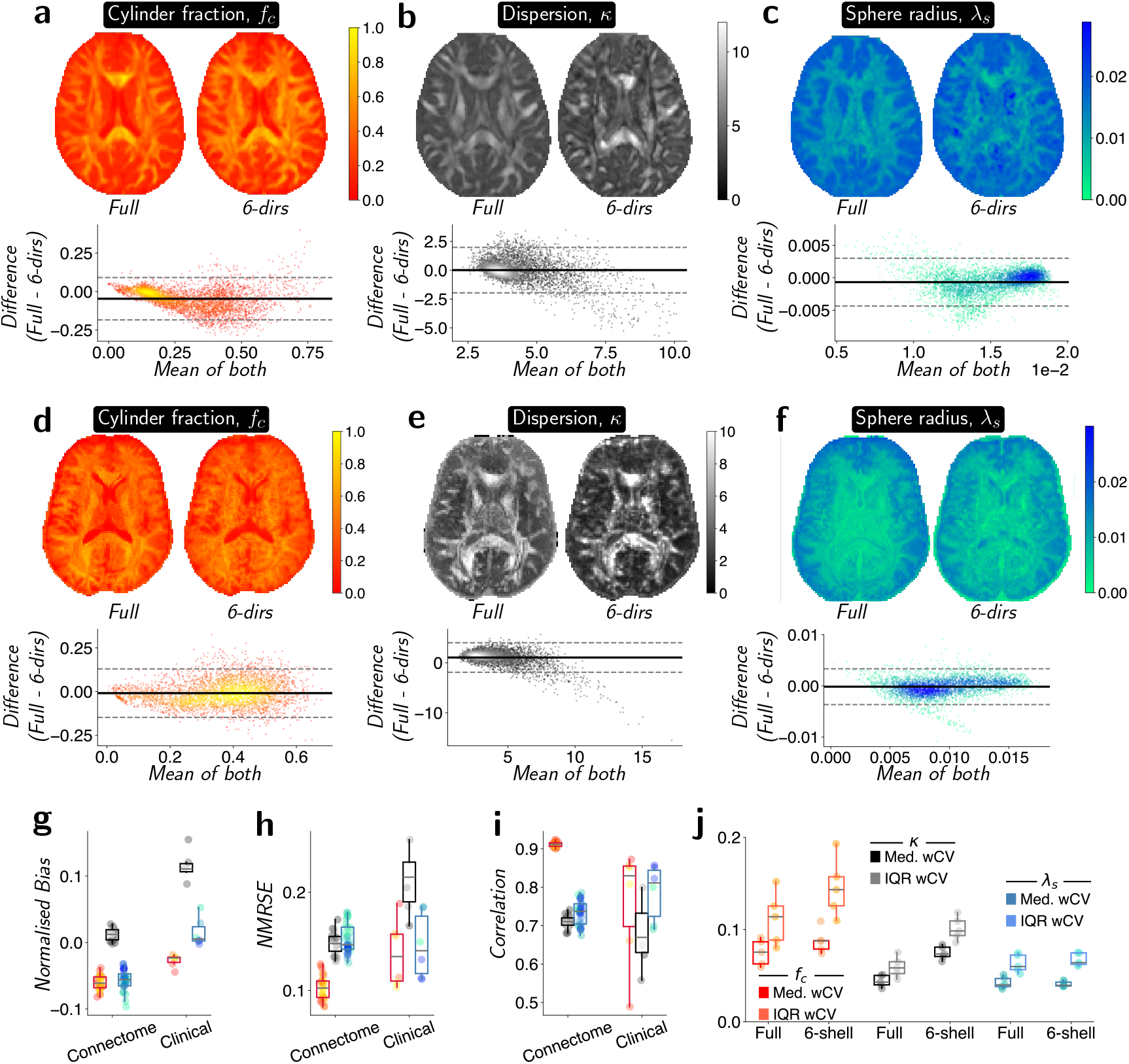
With SBI, a reduced set of 6 acquisitions per b-shell still enables insight into brain microstructure in vivo. **a-c)** Medial axial slices from an example of the high-performance research platform dataset (Garcia-Hernandez et al., 2022) showing three model parameters that have been linked to microglia remodeling, *f*_*c*_, κ and λ_*s*_. In each case, left shows the full acquisition fit (273 acquisitions) while right shows the 6 acquisition per b-shell fit (39 acquisitions). Below, we show a Bland-Altman plot comparing the two for each parameter. Example coronal and sagittal slices can be seen in Fig. S4. **d-f)** Similar to a-c, but the analysis is performed for an individual under the hospital setting clinical acquisition scheme. Here the full dataset represents a 24-direction per shell acquisition as described in the methods. Example coronal and sagittal slices can be seen in Fig. S4. **g-i)** Bias, Normalised root mean squared error (NMRSE) and correlation of parameters obtained for voxels with the full and 6-direction acquisition set for the high-performance research platform (HPRP) (*N* = 29) and hospital setting clinical dataset respectively (*N* = 6). Red, black and blue refer to *f*_*c*_, κ and λ_*s*_, respectively. **j)** Using the test-retest sessions of Garcia-Hernandez et al. (2022), median and the IQR of the wCV are calculated for the different parameters under the different acquisition schemes.

To quantitatively evaluate agreement between the full and reduced protocols across all subjects, we computed the normalised bias, NRMSE, and Pearson correlation coefficient (Fig. 2g–i). Among the evaluated parameters, *f*_*c*_ (red) demonstrated the strongest agreement between protocols across all metrics, whereas κ (black) and λ_*s*_ (blue) showed comparatively lower agreement. For example, in the high-performance research platform dataset, κ exhibited good voxel-wise correspondence between the full and reduced acquisitions, with a Pearson correlation coefficient of 0.71 and an NRMSE of 0.15. Overall, these findings indicate that the reduced protocol preserves the major spatial patterns observed in the full acquisition while introducing moderate local voxel-wise variability. Importantly, normalised bias remained low (1.2%), indicating minimal systematic over- or underestimation between protocols (Fig. 2g).

As expected, agreement was somewhat lower in the hospital setting clinical dataset. For example, κ, which exhibited the lowest overall agreement, showed a normalised bias of approximately 10% and an NRMSE of approximately 0.22. Despite these differences, the principal anatomical contrast patterns observed in the extended acquisition protocol remained evident in the reduced acquisition, suggesting that relative differences between tissue regions are largely preserved even when absolute parameter estimates differ modestly.

In the test–retest analysis (see Fig. 2j), the cylinder fraction exhibited the highest within-subject coefficient of variation (wCV) for both the full and reduced acquisition protocols. However, the strong agreement observed between the two protocols in the across-subject analyses suggests that this variability is unlikely to arise primarily from differences between the fitting approaches and is more consistent with limited reproducibility of the underlying measurement.

#### 3.2.1. Validation, stability and robustness of diffusion-derived microstructural metrics

To assess the regional and inter-subject stability of the extracted microstructural parameters (κ, *f*_*c*_, and λ_*s*_) extracted from the optimised short protocol and to compare them with standard tensor-based metrics (MD and FA) extracted from a traditional multi-shell HARDI acquisition, we computed the coefficient of variation (CoV) across macro-regions and across individuals (averaged across regions) in Fig. 3a and Fig. 3b, respectively. Overall, regional and inter-subject variability were comparable across FA, MD, κ, and *f*_*c*_. In contrast, λ_*s*_ exhibited greater variability, with regional CoV values ranging from 5.6% to 16.8%. Numerically, the highest CoV values were observed in the cerebellum, basal ganglia, hippocampal formation, and amygdala. This pattern may reflect greater biological heterogeneity in the size and composition of soma-rich tissue populations, although increased estimation uncertainty associated with sphere-radius inference may also contribute. A similar trend was observed for inter-subject variability.

**Figure 3:**
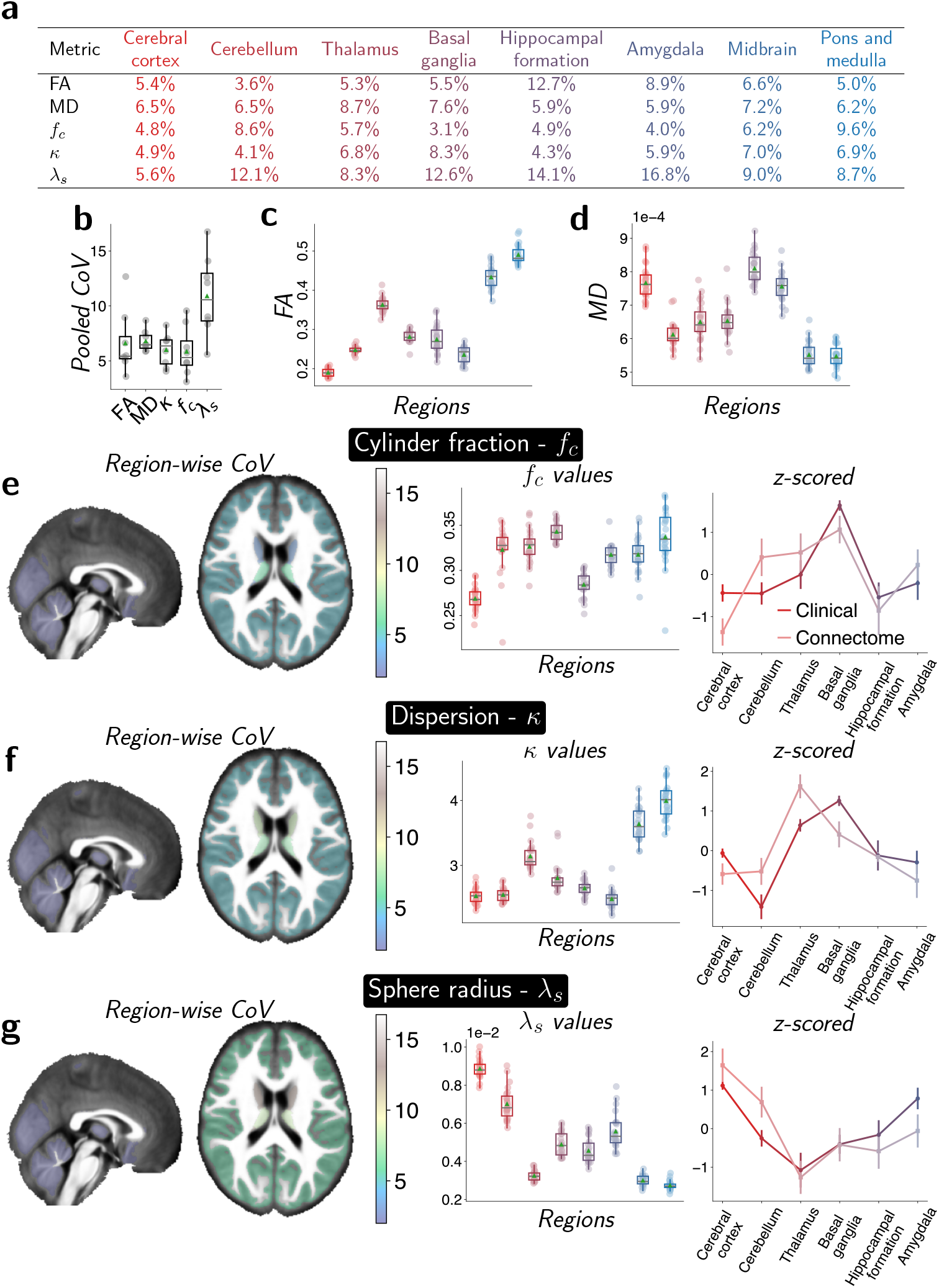
Diffusion-derived microstructural metrics show stable regional distributions and low inter-subject variability. **a)** Table of CoV of the selected model parameters across the macro-regions. **b)** Mean coefficient of variation (CoV) across macro-regions for selected model parameters and metrics. **c-d)** Regional distributions of FA (c) and MD (d) values across macro-regions. **e-g)** CoV of *f*_*c*_ (e), κ (f) and λ_*s*_ (g) projected onto the selected macro-regions. Parameter values across these regions are shown in the middle column, while a *z*-scored version (across regions, darker colors) is compared against values calculated from the high-performance research platform (HPRP) dataset (across regions, lighter colors) in the right column. Pooled CoV and parameter values boxplots represent median values, with boxes showing IQR and whiskers extending to 1.5 times the IQR. Z-scored plots show mean values with error bars showing standard error of the mean.

To formally evaluate these differences, a two-way ANOVA revealed a significant main effect of diffusion metric on CoV values 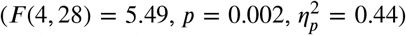. Post hoc Tukey tests showed that λ_*s*_ exhibited significantly greater variability than all other metrics (all *p*< 0.02), whereas FA, MD, κ, and *f*_*c*_ did not differ from one another (all *p* > 0.93). No significant main effect of macro-region was observed 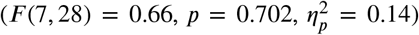, indicating that variability did not differ systematically across brain regions. Across macro-regions, diffusion metrics showed generally stable regional distributions together with low inter-subject variability.

We next examined the regional distribution of the diffusion-derived microstructural metrics by averaging parameter values within each macro-region (Fig. 3c, d and the middle columns of Fig. 3e-g). Distinct spatial patterns were observed across metrics. The cerebral cortex exhibited the highest MD values and among the lowest FA values. Similarly, *f*_*c*_ and κ were generally reduced in the cerebral cortex relative to subcortical structures. More specifically, *f*_*c*_ was highest in the basal ganglia and pons–medulla, whereas κ reached its largest values in the midbrain and pons–medulla. In contrast, λ_*s*_ exhibited a distinct spatial distribution, with the highest values observed in the cerebral cortex and the lowest values in the thalamus. Together, these findings indicate that the proposed diffusion-derived metrics capture systematic regional differences in brain microstructure.

To further evaluate these regional patterns, we compared the parameter distributions with those obtained from the high-performance research platform dataset of Garcia-Hernandez et al. (2022) (Fig. 3e–g). To facilitate direct comparison, parameter values from both datasets were *z*-scored across regions. Although some differences were observed, particularly in the cerebellum and thalamus for *f*_*c*_ and κ, the overall regional trends were largely preserved across datasets. These findings suggest that the clinically feasible acquisition protocol captures similar large-scale microstructural organisation to that observed using the high-gradient high-performance research platform acquisition.

#### 3.2.2. Spatial association between diffusion-derived microstructural metrics and microglial transcriptional signatures

We next investigated whether the simplified multi-compartment model and SBI fitting procedure retained the sensitivity to inflammation-associated microstructural alterations previously reported by Garcia-Hernandez et al. (2022). To this end, the model was applied to five rats from the same study, in which one hemisphere received a saline injection and the contralateral hemisphere received LPS to induce a localized inflammatory response (Fig. 4a). Corresponding histological analyses confirmed marked microglial alterations within the lesion regions (Fig. 4b).

**Figure 4:**
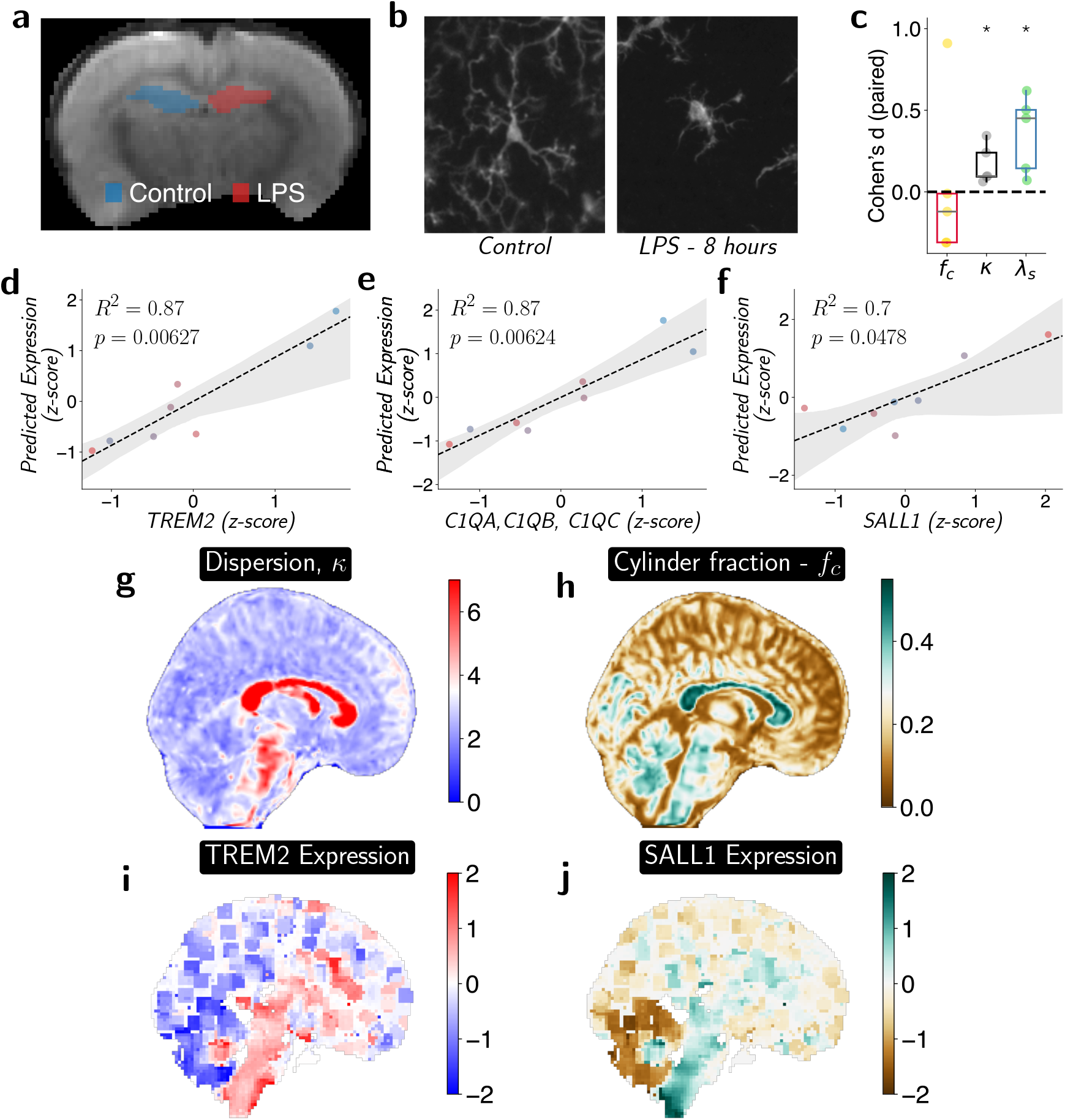
Diffusion-derived microstructural metrics exhibit spatial correspondence with microglial transcriptional signatures. **a)** Representative rat brain with the saline-treated region highlighted in blue and the LPS-treated region highlighted in red. **b)** Representative microglia obtained from Iba-1 histology comparing LPS-and saline-injected tissue. Adapted from Garcia-Hernandez et al. (2022). **c)** Effect size of the two different injection sites (LPS - control) compared for three parameters that were found to be significant in Garcia-Hernandez et al. (2022) (*N* = 5). **d-f)** Multiple regression models relating diffusion-derived microstructural metrics, κ and fraction of cylinders, to HBCA-derived transcriptional measures across eight macro-regions (n = 8). Each point represents one macro-region. Solid lines indicate the fitted relationship between observed and predicted values, and shaded areas represent 95% bootstrapped confidence intervals. Panels correspond to the TREM2 expression d), complement-associated microglial module e) and SALL1 expression f). Model statistics (*R*^2^ and *F*-test *p*-values) are shown in each panel. **g-j)** Upper line: cohort-averaged κ and *f*_*c*_; lower line: TREM2 and SALL1 gene expression maps from Neurosynth, displaying human genes contained in the Allen Human Brain Atlas microarray dataset (Shen et al., 2012). Example axial and coronal slices can be seen in Fig. **??**.

As shown in Fig. 4c, lesions exhibited the same directional changes in the diffusion-derived metrics as reported previously, including increased κ values, indicative of reduced dispersion, and increased λ_*s*_. The concordance between the present results and the original study suggests that the simplified model preserves sensitivity to microstructural changes associated with experimentally induced neuroinflammation.

To evaluate whether diffusion-derived microstructural metrics exhibit relationships with microglial transcriptional signatures in the human brain, we performed multiple linear regression analyses across eight macro-regions using κ and *f*_*c*_ as joint predictors of Human Brain Cell Atlas (HBCA)-derived gene expression profiles.

Microglial markers included the complement-associated module (C1QA, C1QB, and C1QC) (Boche and Gordon, 2022), TREM2 (Colonna and Wang, 2016), SALL1 (Buttgereit et al., 2016), TGFB1 (Sobue et al., 2021), P2RY12, and TMEM119 (Butovsky and Weiner, 2018), representing genes associated with homeostatic, pruning-related, and microglial identity programs. As non-microglial controls, we included APOE and AQP4, which are predominantly expressed by astrocytes (Matejuk and Ransohoff, 2020; Peng et al., 2023).

For the complement-associated module, the regression model was significant (*F* (2, 5) = 16.51, *p* = 0.0063) and explained a large proportion of regional variance (*R*^2^ = 0.869, adjusted *R*^2^ = 0.816; Fig. 4e). Predicted and observed expression values were strongly correlated (*r* = 0.932, *p* = 0.00075), indicating that the regional distribution of complement-associated gene expression closely corresponded to the diffusion-derived microstructural metrics. Among the predictors, κ showed the dominant positive association (β_κ_ = 0.819), whereas *f*_*c*_ exhibited a weaker contribution 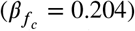. Bootstrap analysis confirmed the robustness of the κ effect (*p*_boot_ = 0.0056), while *f*_*c*_ did not contribute independently (*p*_boot_ = 0.3104).

A highly similar pattern was observed for TREM2 (Fig. 4d). The regression model explained a comparable proportion of variance (*R*^2^ = 0.869, adjusted *R*^2^ = 0.816; *F* (2, 5) = 16.55, *p* = 0.0062), and predicted values closely matched observed expression levels (*r* = 0.932, *p* = 0.00074). As with the complement-associated module, κ was the dominant predictor (β_κ_ = 0.863), whereas *f*_*c*_ contributed only weakly 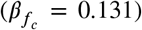. Bootstrap analysis yielded a borderline result for the κ effect (*p*_boot_ = 0.0544). Notably, the regional distribution of TREM2 closely mirrored that of the complement-associated module, suggesting that both transcriptional signatures share a common spatial organisation and microstructural correlate captured primarily by κ.

In contrast, SALL1 exhibited a distinct relationship with the diffusion-derived metrics (Fig. 4f). Although the regression model remained significant (*F* (2, 5) = 5.94, *p* = 0.0478, *R*^2^ = 0.704, adjusted *R*^2^ = 0.585), the dominant association was a negative effect of 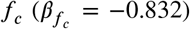, whereas κ showed little contribution (β_κ_ = −0.015). Thus, although SALL1 also displayed a spatial distribution that was captured by the diffusion-derived metrics, its underlying microstructural associations differed from those observed for the complement-associated module and TREM2.

To further assess the selectivity of these associations, additional microglia-enriched markers were examined. TGFB1 showed a trend-level association (*F* (2, 5) = 4.65, *p* = 0.0723, *R*^2^ = 0.650; Supplementary Fig. S2a), indicating partial spatial correspondence with the diffusion-derived metrics. In contrast, TMEM119 (*F* (2, 5) = 2.16, *p* = 0.211, *R*^2^ = 0.464) and P2RY12 (*F* (2, 5) = 1.17, *p* = 0.383, *R*^2^ = 0.319) did not exhibit significant spatial associations (Supplementary Fig. S2b,c). These findings indicate that spatial correspondence with the diffusion-derived metrics is not uniformly observed across all microglial-enriched genes.

As a further control, regression analyses were performed using the astrocyte-associated markers APOE (*F* (2, 5) = 0.85, *p* = 0.481, *R*^2^ = 0.254) and AQP4 (*F* (2, 5) = 0.53, *p* = 0.618, *R*^2^ = 0.175). Although both genes exhibited regional variation within the HBCA atlas, neither showed significant correspondence with the diffusion-derived metrics (Supplementary Fig. S2d,e). The absence of comparable associations for these non-microglial markers supports the interpretation that the observed relationships are preferentially linked to specific microglial transcriptional programs rather than reflecting generic large-scale regional gradients. Full regression statistics are provided in Supplementary Table S4.

To visualise and compare the spatial distribution of the inferred transcriptional signatures with the relevant dMRI markers, in Fig. 4g-j we report cohort-averaged κ and fraction of cylinders maps along with the TREM2 and SALL1 gene expression maps from Neurosynth.org, which are derived from human gene expression data in the Allen Human Brain Atlas. Consistent with the regression analyses, the spatial distribution of TREM2 expression closely resembled that of κ, whereas the SALL1 expression map exhibited a pattern more similar to the cylinder fraction. These qualitative observations further support the associations identified in the transcriptomic analyses.

### 3.3. Exploratory application of the NEXI model

To demonstrate that our approach is applicable to models and time-dependent quantities beyond the one introduced in this study, we additionally fit the NEXI model introduced by Jelescu et al. (2022) within our acquisition and SBI framework. We mirror the work of Uhl et al. (2025), who likewise did not satisfy the short-gradient-pulse approximation, but emphasize that our acquisition protocol uses significantly fewer b-shells and directions, resulting in a total scan time that is roughly one-third shorter.

We first evaluated the NEXI fitting framework *in silico* by applying the trained SBI network to unseen signals generated using the same simulator (Fig. 5a–d). We considered four NEXI parameters: the cylinder fraction, *f*; the exchange time, *t*_ex_; and the intracellular and extracellular diffusivities, *D*_*i*_ and *D*_*e*_, respectively. The estimated *f*, *D*_*i*_, and *D*_*e*_ showed good agreement with their ground-truth values, with correlations of 0.991, 0.85, and 0.92, respectively. In contrast, *t*_ex_ was less accurately recovered, with a correlation of approximately 0.58. This reduced sensitivity to exchange likely reflects the limited exchange sensitivity of the acquisition, particularly given the relatively low diffusion-encoding gradient strengths.

**Figure 5:**
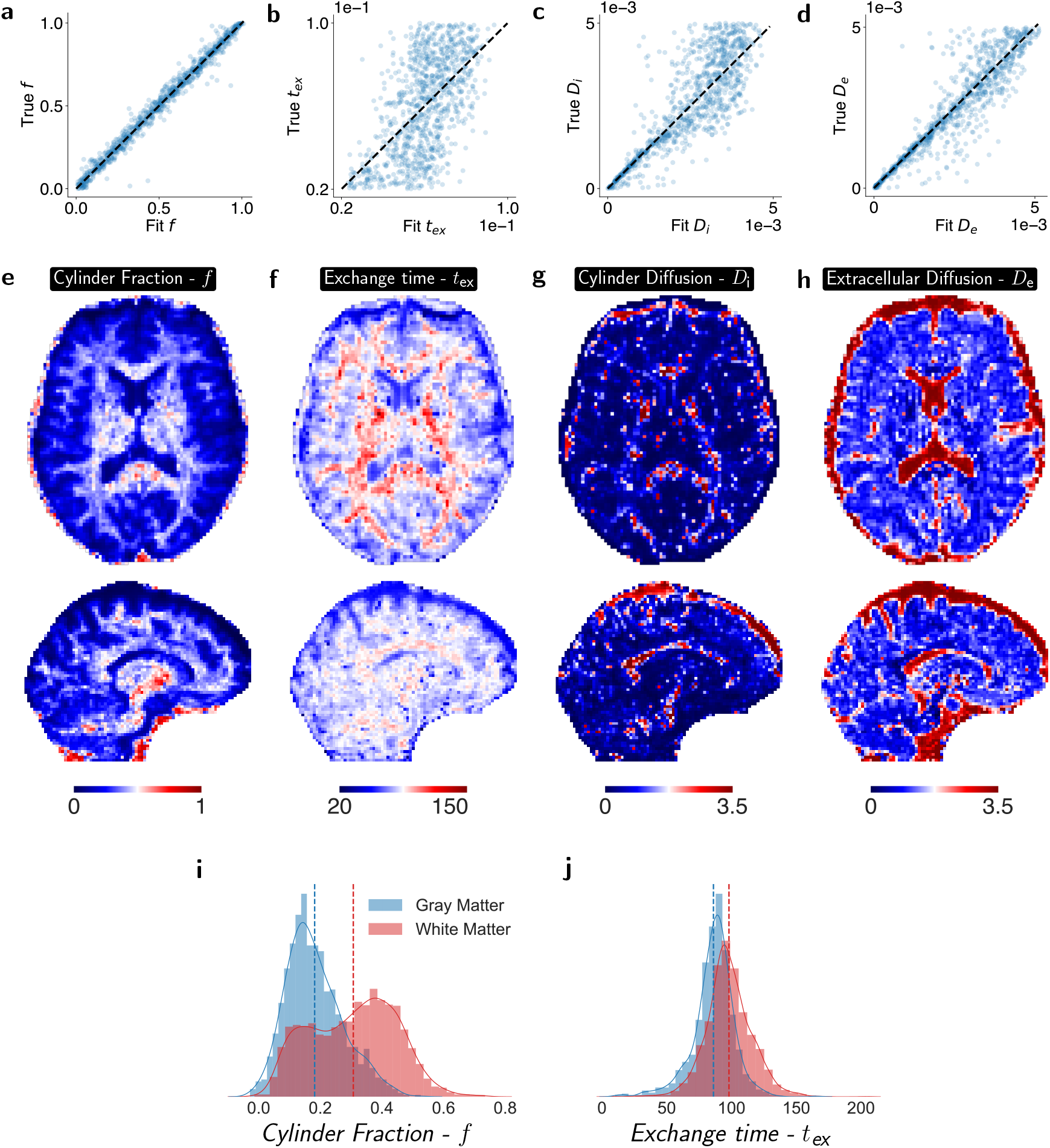
*In silico* and *in vivo* evaluation of the NEXI model using our acquisition scheme. **a-d)** Comparison of NEXI parameters estimated using the SBI pipeline against the ground-truth parameters for 1000 randomly sampled test signals. **e-h)** NEXI parameter maps obtained by applying the same SBI pipeline to one participant from our clinical cohort using the 13-acquisition protocol. An example axial and sagittal slice is shown for each of the four NEXI parameters. **i-j)** Distributions of the NEXI cylinder fraction, *f*, and exchange time, *t*_ex_, in white (red) and gray (blue) matter for the same participant. Dashed vertical lines indicate the mean value for each tissue class.

Next, we applied the NEXI SBI fitting pipeline *in vivo* to one participant who underwent the shortened acquisition protocol. Example medial-axis parameter maps are shown in Fig. 5e–h. The estimated cylinder fraction, *f*, was spatially well resolved and exhibited patterns similar to those reported by Uhl et al. (2025). The *t*_ex_ map was noisier, but the estimated values were broadly consistent with those previously reported. In contrast, the *D*_*i*_ and *D*_*e*_ maps were poorly resolved. This is also consistent with Uhl et al. (2025), who reported substantial variability in *D*_*i*_, with estimates frequently reaching the upper fitting bound. Notably, because *D*_*i*_ and *D*_*e*_ were well identified in the matched *in silico* experiment, their poor in-vivo recovery is unlikely to arise solely from limitations of the SBI estimator. Instead, it may reflect model mismatch and reduced parameter identifiability in vivo, potentially arising from the finite gradient pulse duration, comparatively low gradient strengths, and tissue features not captured by the NEXI model.

Finally, we examined whether the NEXI-derived *f* and *t*_ex_ differed between gray and white matter by plotting their respective distributions (Fig. 5i,j). White matter exhibited a higher cylinder fraction, consistent with its greater abundance of coherently oriented neurites, as well as longer estimated exchange times, which reflects differences in membrane permeability and myelination between gray and white matter and are qualitatively consistent with previous observations (Uhl et al., 2025). Together, these results demonstrate that, even without optimizing the acquisition specifically for NEXI, the proposed acquisition and SBI framework can be extended to other advanced diffusion MRI models and used to probe time-dependent quantities such as inter-compartmental exchange.

## 4. Discussion

This study establishes the feasibility of clinically practical diffusion MRI that simultaneously interrogates diffusion weighting and diffusion time dependence, while maintaining an acquisition time compatible with routine clinical workflows. The key enabling factors are two: an optimised acquisition design, and the recently proposed simulation-based inference framework for model fitting (Eggl and De Santis, 2026), which together maximize information content and substantially reduce the number of measurements required. As a result, advanced microstructural imaging strategies that have traditionally been confined to specialised research environments can be implemented on clinically relevant timescales, expanding their potential utility for the diagnosis and monitoring of neurological diseases.

Given the increased sensitivity and specificity of non-tensor diffusion MRI approaches relative to conventional diffusion tensor imaging (De Santis et al., 2014), substantial efforts have been devoted to translating advanced microstructural models into clinical practice. The neurite orientation dispersion and density imaging (NODDI) model was designed from the outset with clinical feasibility in mind and can be robustly estimated using a relatively simple two-shell acquisition protocol (Timmers et al., 2016). More recently, the soma and neurite density imaging (SANDI) model (Palombo et al., 2020) has extended this framework by providing additional measures related to cellularity and soma density. Despite its greater model complexity, SANDI has been shown to be feasible on hospital setting clinical 3T scanners using a 10-minute acquisition (Schiavi et al., 2023) or 15-minute (Barakovic et al., 2024) protocol, albeit under the assumption of negligible inter-compartmental water exchange.

Here, we successfully extend this concept to acquisition protocols that additionally probe the diffusion-time dimension. Models relying on varying diffusion times have originally been introduced to either estimate the physical size of diffusion compartments as in AxCaliber (Assaf et al., 2008), by exploiting the fact that the diffusion signal arising from restricted compartments varies as a function of diffusion time, or to overcome the limitations of assuming impermeable compartments including Neurite exchange imaging (NEXI) and Standard Model with Exchange (SMEX), protocols that sample multiple diffusion times to robustly estimate exchange (Jelescu et al., 2022; Olesen et al., 2022). Implementation of time-dependent diffusion MRI has recently been studied in clinical settings. For example, Grussu et al. (2019) performed one of the first clinical multi-diffusion-time studies in the human spinal cord, a tissue characterized by a highly coherent and anisotropic axonal architecture. More recently, Hertanu et al. (2025) and Uhl et al. (2025) implemented multi-diffusion-time acquisitions on clinical MRI systems to investigate exchange effects in gray matter using the NEXI and SMEX frameworks, respectively. These studies demonstrate the growing feasibility of advanced time-dependent diffusion imaging on clinical hardware and provide an important foundation for the present work. However, these studies, primarily designed to investigate exchange-related microstructural effects, employed substantially longer acquisition protocols (typically exceeding 25 min). Here, we substantially lower thetime requirement, proposing an implementation that is compatible with clinical needs.

Besides protocols that exploit diffusion-time dependence using conventional spin-echo diffusion sequences, alternative approaches have been proposed based on non-conventional diffusion encoding schemes, such as double diffusion encoding (Yang et al., 2018; Henriques et al., 2021) and arbitrary gradient waveform encoding (Westin et al., 2016). These methods may provide increased sensitivity to specific microstructural features and enable the estimation of parameters like the microscopic anisotropy, a biomarker that has been linked to features such as cell shape and neurite geometry (Ianus et al., 2021). However, the aim of the present work was not to develop or evaluate alternative encoding schemes, but rather to optimise a standard diffusion MRI acquisition that can be readily implemented in routine clinical settings, given that the clinical translation of non-standard diffusion sequences remains particularly challenging due to increased implementation complexity, hardware requirements, and limited availability on commercial scanners.

Our parameter estimation performance analysis demonstrated that the proposed protocol achieves levels of accuracy comparable to those typically reported for standard tensor-based diffusion MRI metrics. Importantly, these comparisons should be interpreted in light of the fact that the tensor-derived measures were obtained from a conventional highangular-resolution acquisition protocol with a substantially longer scan time than the optimised multi-Δ protocol.

Interestingly, microstructural maps in our cohort exhibited spatially structured associations with microglial transcriptional signatures across the human brain. The strongest relationships were observed for the complement genes (C1QA, C1QB, C1QC) and TREM2, key mediators of synaptic pruning and debris clearance. In the healthy brain, C1Q produced by microglia tags weak or redundant synapses for elimination, whereas TREM2 promotes the detection and clearance of damaged myelin and apoptotic debris (Stevens et al., 2007; Presumey et al., 2017; Colonna and Wang, 2016). Their association with κ therefore suggests sensitivity to microglia-driven remodelling of tissue architecture and extracellular organisation, rather than simply reflecting cellular density. A distinct pattern was observed for SALL1, a master regulator of homeostatic microglial identity (Buttgereit et al., 2016), whose enrichment in cortical and hippocampal regions is consistent with known regional heterogeneity of microglial states in the adult human brain (Sankowski et al., 2019). By contrast, no significant associations were detected for the astrocytic markers APOE and AQP4, supporting the specificity of the observed relationships to microglial biology. Taken together, these findings highlight the potential of our framework for non-invasive characterisation of cellular architecture in vivo and contribute to the growing field of imaging transcriptomics, in which MRI-derived spatial maps are linked to gene expression atlases to infer their underlying cellular and molecular substrates (Arnatkeviciūtė et al., 2019; Seidlitz et al., 2020).

It is worth mentioning that while a large part of the present work focused on a single microstructural model, the acquisition scheme is model-agnostic and the data collected can be processed with alternative formulations, including models that explicitly account for water exchange between compartments, as demonstrated by our exploratory analysis of the NEXI model. This model was recently applied in a clinical setting similar to the one considered here (Uhl et al., 2025). As in that study, however, our acquisition does not necessarily satisfy the short-gradient-pulse approximation (δ ≪ Δ) required for the analytical solution derived by Jelescu et al. (2022). Importantly, our primary aim was to demonstrate the generalizability of the proposed acquisition and SBI framework, showing that it can be extended to time-dependent processes beyond microstructural size, with exchanging estimation serving as a proof of concept at this point. Future work could include using more advanced models, such as SMEX (Olesen et al., 2022) that relaxes the short-gradient pulse approximation but require numerical solution of an ordinary differential equation to simulate the diffusion-weighted signal. Moreover, future extensions could incorporate joint diffusion–relaxometry acquisitions, enabling the simultaneous estimation of diffusion and relaxation parameters (De Santis et al., 2016a; Slator et al., 2021; Barakovic et al., 2023).

To conclude, these findings suggest that the multi-Δ framework captures biologically meaningful aspects of tissue microstructure using an acquisition protocol that is compatible with routine clinical imaging. This positions the framework as a promising non-invasive tool for advanced microstructural characterization in vivo. Having established feasibility and reproducibility in healthy participants, the framework is now well positioned for evaluation in clinical cohorts, where it may support the early detection, stratification, and monitoring of neurological disorders in which cell-specific alterations play a central role.

## Data Availability

The code for neural network training, model fitting, and inference is publicly available at GitHub. The study also uses publicly available animal and human MRI data from Garcia-Hernandez et al. (2022). Newly acquired datasets are not publicly available at the time of submission but will be made publicly available upon publication.

https://github.com/meggl23/ClinicalAdvancedDwMRI

https://www.science.org/doi/10.1126/sciadv.abq2923

## Acknowledgements

We acknowledge the support of the Institute of Neuroscience, CSIC-UMH. We thank Prof. Israel Dudkevich for his institutional support of this project, including oversight of the regulatory and ethical framework that enabled participant recruitment and the successful conduct of the study.

This research was funded by “la Caixa” Foundation with Project-ID LCF/BQ/PI23/11970039 (MFE). This study was supported by Nehemia Rubin Excellence in Biomedical Research–the TELEM Program research grant (AbiL). SDS was supported by the Spanish Ministerio de Ciencia e Innovación, Agencia Estatal de Investigación (PID2025-175422OB-I00 and CNS2023-14488), by the Programs for Centres of Excellence in R&D Severo Ochoa (CEX2021-001165-S), by the Generalitat Valenciana through a Subvencion para la contratación de investigadoras e investigadores doctores de excelencia 2021 (CIDEGENT/2021/015), and by the Pasqual Maragall Foundation under the Pasqual Maragall Researchers Programme (PMRP) (Grant Agreement Number 2023-1296).

We thank the Cardiff University Brain Research Imaging Centre (CUBRIC) and Prof. Derek Jones for sharing the re-test data of the high-performance MRI dataset (Garcia-Hernandez et al., 2022).

## 5. Declaration of Generative AI Use

During the preparation of this manuscript, the authors used ChatGPT (GPT-5.6 Sol, OpenAI) for reviewing, copyediting, and improving the clarity of the text. The scientific content, interpretation of results, and initial and final drafting were performed and verified by the authors. All references and citations were selected and independently verified by the authors. The authors take full responsibility for the content of the manuscript.

## CRediT authorship contribution statement

**Anat Leibovici:** Conceptualization, Methodology, Formal analysis, Software, Data Curation, Validation, Investigation, Writing - Original Draft, Writing - Review & Editing, Visualization. **Elena Espinós Soler:** Methodology, Software, Data Curation, Validation, Writing - Original Draft. **David Mesika:** Methodology, Software. **Galia Tsarfaty:** Validation, Formal analysis, Data Curation. **Abigail Livny:** Conceptualization, Resources,Writing - Original Draft,, Writing - Review & Editing, Supervision, Project administration, Funding acquisition. **Silvia De Santis:** Conceptualization, Methodology, Resources, Writing - Original Draft, Writing - Review & Editing, Supervision, Project administration, Funding acquisition. **Maximilian F. Eggl:** Conceptualization, Methodology, Writing - Original Draft, Writing - Review & Editing, Supervision, Funding acquisition, Visualization.

