## Supplemental information for "A Time-Dependent Diffusion MRI Framework for Clinical Characterisation of Human Brain Cellular Architecture"

### A. Participant Gender/Age information

| Group | N | Mean | Median | SD | Min | Max |
|---|---|---|---|---|---|---|
| Male | 12 | 30.83 | 30.00 | 3.30 | 25.00 | 38.00 |
| Female | 11 | 30.45 | 30.00 | 4.86 | 22.00 | 39.00 |

**Supplementary Table S1**
*Age distribution of the study cohort by sex.* Values are reported in years as mean ± s.d., with median and range (minimum–maximum), for males ($n = 12$) and females ($n = 11$).

### B. Additional MRI analysis

#### B.1. Quality Control and Intensity Outlier Detection

Quality assurance comprised two independent pipelines addressing distinct failure modes, applied prior to statistical analysis.

**Motion quality control:** Subject-level motion estimates were derived from FSL eddy output parameters, extracted from the `-eddyqc_all` output directory generated during preprocessing. For each participant, the mean, maximum, and standard deviation of volume-to-volume displacement were computed across all 39 volumes of the concatenated dataset. Participants with mean displacement exceeding 0.4 mm were excluded, consistent with thresholds applied in prior dMRI quality control studies (Bastiani et al., 2019). $Z$-score summaries across the cohort are provided for reference in Supplementary Table S2.

**Intensity outlier detection:** Separately, subject-level distributions of the three diffusion-derived microstructural parameter maps, $\kappa$, fraction of cylinders, $f_c$ , and average radius of the sphere $\lambda_s$, were assessed to identify gross parameter estimation failures, such as those arising from signal dropout, reconstruction instability, or severe co-registration errors. For each metric and each participant, two summary statistics were computed across all voxels within the whole-brain diffusion mask: the subject-wise spatial mean, reflecting the overall signal level, and the subject-wise spatial standard deviation, reflecting within-subject map variability. Both statistics were standardised using $z$-scores across the cohort. A threshold of $|z| > 2.5$ was applied to flag potential outliers, evaluated independently for mean and variability estimates of each metric. This threshold corresponds to a conservative criterion that, in a normally distributed sample of n = 24, yields a family-wise expected false-positive rate below 5% across the six $z$-score comparisons performed (Grubbs, 1969). Subject-level summary statistics and $z$-scores for all metrics are reported in Supplementary Table S3.

**Visual quality control:** All diffusion parameter maps and their overlay on the corresponding T1-weighted anatomical images were visually inspected by an experienced researcher for evidence of signal dropout, EPI-related distortion artefacts, reconstruction failures, and gross co-registration errors between diffusion and anatomical space. Inspection was performed independently of the intensity $z$-score flags.

#### B.2. Brain Parcellation and Regional Signal Extraction

High-resolution T1-weighted images were processed using the FreeSurfer recon-all pipeline (Fischl, 2012) to generate participant-specific cortical and subcortical parcellations. Cortical regions were defined according to the Desikan–Killiany atlas (Desikan et al., 2006). For each participant, anatomical parcellations were co-registered to diffusion space, and regional mean diffusion values were extracted using in-house Python (version 3.8) scripts across cortical, subcortical, cerebellar, and brainstem structures for all diffusion metrics, including fractional anisotropy (FA), mean diffusivity (MD), dispersion parameter $\kappa$, fraction of cylinders, $f_c$, and average radius of the sphere, $\lambda_s$, compartment. For cross-modal regression analyses, regional values of $\kappa$ and $f_c$ were aggregated into eight macro-regions: cerebral cortex, cerebellum, thalamus, basal ganglia, hippocampal formation, amygdala, midbrain, and pons–medulla. Macro-regions were defined by bilaterally averaging corresponding left and right hemisphere structures using mean-wise aggregation of regional diffusion values. The macro-regional scheme was aligned with the anatomical definitions provided in the Human Brain Cell Atlas (HBCA) to ensure spatial correspondence between MRI-derived regional metrics and transcriptomic data for subsequent cross-modal analyses.

#### B.3. Inter-subject variability and validation of diffusion-derived microstructural metric

Inter-subject variability of diffusion metrics was quantified using the coefficient of variation (CoV; SD/mean × 100), computed across participants for each diffusion metric within the eight predefined macro-regions. Macro-region–wise

CoV values were averaged to obtain a mean CoV per metric and were also examined individually to compare variability across brain structures. To assess consistency of parameter estimates with the original implementation of the multi-$\Delta$ stick-and-ball framework (Garcia-Hernandez et al., 2022), distributions of the $\kappa$ and $f_c$ in the hippocampal formation were compared with values reported in the human pilot dataset (n = 6) described in that study. Group means were compared and variability was summarised using the standard error of the mean (SEM), calculated as SD/$\sqrt{n}$, where $n$ denotes the sample size. As the two datasets were obtained under different acquisition conditions and parameter ranges, values were normalised. Differences in inter-subject variability across diffusion metrics and brain regions were evaluated using a two-way analysis of variance (ANOVA) with diffusion metric (FA, MD, $\kappa$, $f_c$, and $\lambda_s$) and macro-region (eight regions) as factors. The dependent variable was the CoV calculated across participants for each region and metric. Effect sizes are reported as partial eta squared ($\eta_p^2$). Post-hoc pairwise comparisons were performed using Tukey's honestly significant difference (HSD) test.

#### B.4. Imaging-Derived Transcriptional Mapping and Spatial Visualisation

To generate spatial representations of transcriptional signatures derived from cross-modal regression analyses, $\kappa$ and $f_c$ were combined using fixed weights corresponding to the standardised regression coefficients obtained at the macro-regional level. For each transcriptional signature (complement-associated module/TREM2 and SALL1), the respective standardised beta coefficients linking $\kappa$ and $f_c$ to the HBCA measure were applied uniformly to parcel-wise diffusion values across the brain. This procedure yielded imaging-derived transcriptional maps reflecting the spatial distribution implied by the regression model. Cohort-averaged parameter maps ($\kappa$ and $f_c$) were projected onto the Desikan atlas to obtain a visual representation of spatial distribution and then compared against gene expression maps extracted from Neurosynth.org and the Allen brain atlas (Shen et al., 2012). These maps were generated for descriptive visualisation only. No inferential statistical testing was performed.

#### B.5. Secondary analyses: sex-related variation in diffusion-derived microstructural metrics

Sex-related effects on $\kappa$ and $f_c$ were tested using ANCOVA with age as a covariate, with false discovery rate (FDR) correction applied across the eight macro-regions. Analyses were performed for $\kappa$, $f_c$, the complement-associated module–derived map, TREM2-derived map, and SALL1-derived map. No significant sex differences were observed for any metric after FDR correction across regions (see Supplementary table S5).

| Subject | Mean Motion | Max Motion | Std Motion | N Volumes | Volume Z-score | Max Motion Z-score | QC Status |
|---|---|---|---|---|---|---|---|
| #01 | 0.443 | 2.872 | 0.647 | 39 | 2.987 | 2.388 | Excluded |
| #02 | 0.224 | 2.758 | 0.446 | 39 | 0.810 | 2.251 | - |
| #03 | 0.328 | 2.403 | 0.517 | 39 | 1.844 | 1.823 | - |
| #04 | 0.299 | 2.327 | 0.459 | 39 | 1.552 | 1.731 | - |
| #05 | 0.028 | 0.156 | 0.039 | 39 | -1.140 | -0.887 | - |
| #06 | 0.033 | 0.227 | 0.064 | 39 | -1.090 | -0.801 | - |
| #07 | 0.248 | 1.106 | 0.270 | 39 | 1.045 | 0.258 | - |
| #08 | 0.042 | 0.179 | 0.064 | 39 | -0.996 | -0.859 | - |
| #09 | 0.044 | 0.334 | 0.078 | 39 | -0.978 | -0.672 | - |
| #10 | 0.067 | 0.196 | 0.047 | 39 | -0.751 | -0.839 | - |
| #11 | 0.060 | 0.289 | 0.080 | 39 | -0.818 | -0.726 | - |
| #12 | 0.069 | 0.225 | 0.061 | 39 | -0.728 | -0.804 | - |
| #13 | 0.078 | 0.303 | 0.073 | 39 | -0.638 | -0.710 | - |
| #14 | 0.092 | 0.362 | 0.096 | 39 | -0.501 | -0.639 | - |
| #15 | 0.093 | 0.433 | 0.086 | 39 | -0.496 | -0.553 | - |
| #16 | 0.120 | 0.444 | 0.101 | 39 | -0.220 | -0.539 | - |
| #17 | 0.180 | 1.309 | 0.242 | 39 | 0.375 | 0.503 | - |
| #18 | 0.139 | 1.238 | 0.196 | 39 | -0.032 | 0.418 | - |
| #19 | 0.129 | 0.576 | 0.116 | 39 | -0.135 | -0.380 | - |
| #20 | 0.158 | 0.584 | 0.113 | 39 | 0.157 | -0.371 | - |
| #21 | 0.114 | 0.774 | 0.131 | 39 | -0.284 | -0.141 | - |
| #22 | 0.117 | 0.671 | 0.130 | 39 | -0.249 | -0.266 | - |
| #23 | 0.161 | 0.729 | 0.162 | 39 | 0.188 | -0.196 | - |
| #24 | 0.152 | 0.900 | 0.194 | 39 | 0.098 | 0.011 | - |

**Supplementary Table S2**
**Subject-level motion correction quality control for diffusion-derived microstructural metrics.** Participant-wise motion estimates derived from FSL eddy, extracted from the `-eddyqc_all` output generated by `dwifslpreproc` (MRtrix3). Motion correction was applied jointly to the concatenated 39-volume dataset ($\Delta = 27$, 40, and 60 ms acquired sequentially), ensuring all diffusion times share a common motion reference frame. Mean, maximum, and standard deviation of volume-to-volume displacement are reported per participant. Motion parameters were standardised using z-scores across the full cohort (n = 24); participants with $|z| > 2.5$ specifically on the mean motion estimate were flagged for exclusion. One participant (mean motion = 0.443 mm; $z = 2.99$) was excluded, yielding a final analytic sample of n = 23. Among included participants, mean motion was 0.129 mm.

| Subject | $f_c$ Mean | $f_c$ SD | $\kappa$ Mean | $\kappa$ SD | $\lambda_s$ Mean | $\lambda_s$ SD | $f_c$ Mean Z | $f_c$ SD Z | $\kappa$ Mean Z | $\kappa$ SD Z | $\lambda_s$ Mean Z | $\lambda_s$ SD Z |
|---|---|---|---|---|---|---|---|---|---|---|---|---|
| #01 | 0.082 | 0.151 | 0.902 | 1.566 | 0.002 | 0.004 | -0.074 | -0.289 | 0.013 | -0.476 | 0.029 | 0.086 |
| #02 | 0.071 | 0.146 | 0.732 | 1.473 | 0.002 | 0.003 | -1.610 | -1.278 | -1.824 | -1.764 | -0.773 | -0.313 |
| #03 | 0.089 | 0.161 | 0.930 | 1.654 | 0.002 | 0.004 | 0.786 | 1.446 | 0.314 | 0.740 | 0.239 | 0.257 |
| #04 | 0.079 | 0.152 | 0.823 | 1.528 | 0.002 | 0.004 | -0.428 | -0.234 | -0.840 | -1.008 | -0.387 | -0.150 |
| #05 | 0.083 | 0.153 | 0.992 | 1.696 | 0.002 | 0.004 | 0.071 | -0.069 | 0.985 | 1.318 | 0.857 | 0.855 |
| #06 | 0.091 | 0.161 | 0.954 | 1.658 | 0.002 | 0.004 | 1.075 | 1.420 | 0.579 | 0.791 | 0.313 | 0.190 |
| #07 | 0.073 | 0.151 | 0.705 | 1.442 | 0.002 | 0.004 | -1.218 | -0.404 | -2.120 | -2.204 | -0.089 | -0.031 |
| #08 | 0.089 | 0.153 | 0.943 | 1.552 | 0.002 | 0.004 | 0.887 | -0.052 | 0.452 | -0.670 | 0.196 | 0.058 |
| #09 | 0.079 | 0.147 | 0.900 | 1.583 | 0.002 | 0.004 | -0.529 | -1.032 | -0.009 | -0.245 | 0.152 | 0.311 |
| #10 | 0.074 | 0.147 | 0.803 | 1.541 | 0.002 | 0.004 | -1.131 | -1.095 | -1.063 | -0.825 | -0.135 | 0.098 |
| #11 | 0.088 | 0.157 | 0.968 | 1.666 | 0.002 | 0.004 | 0.698 | 0.747 | 0.724 | 0.900 | 0.431 | 0.348 |
| #12 | 0.082 | 0.149 | 0.978 | 1.663 | 0.002 | 0.004 | -0.136 | -0.633 | 0.835 | 0.864 | 0.830 | 0.805 |
| #13 | 0.094 | 0.160 | 1.094 | 1.737 | 0.002 | 0.004 | 1.541 | 1.222 | 2.096 | 1.896 | 0.917 | 0.535 |
| #14 | 0.084 | 0.151 | 0.906 | 1.559 | 0.002 | 0.004 | 0.195 | -0.257 | 0.062 | -0.580 | 0.133 | 0.168 |
| #15 | 0.067 | 0.140 | 0.747 | 1.532 | 0.002 | 0.003 | -2.040 | -2.171 | -1.669 | -0.945 | -0.453 | -0.273 |
| #16 | 0.088 | 0.157 | 1.004 | 1.693 | 0.002 | 0.004 | 0.764 | 0.645 | 1.123 | 1.278 | 0.805 | 0.624 |
| #17 | 0.086 | 0.153 | 1.003 | 1.677 | 0.002 | 0.003 | 0.510 | 0.092 | 1.111 | 1.055 | 0.505 | 0.521 |
| #18 | 0.074 | 0.148 | 0.850 | 1.616 | 0.002 | 0.004 | -1.121 | -0.850 | -0.549 | 0.209 | 0.342 | 0.351 |
| #19 | 0.076 | 0.146 | 0.875 | 1.543 | 0.002 | 0.004 | -0.871 | -1.268 | -0.282 | -0.801 | 0.261 | 0.337 |
| #20 | 0.091 | 0.159 | 0.945 | 1.615 | 0.002 | 0.004 | 1.129 | 1.087 | 0.478 | 0.198 | -0.111 | -0.179 |
| #21 | 0.083 | 0.154 | 0.947 | 1.641 | 0.002 | 0.004 | 0.092 | 0.246 | 0.500 | 0.562 | 0.475 | 0.445 |
| #22 | 0.089 | 0.158 | 0.878 | 1.553 | 0.002 | 0.003 | 0.839 | 0.924 | -0.244 | -0.660 | -0.488 | -0.454 |
| #23 | 0.095 | 0.164 | 0.924 | 1.607 | 0.002 | 0.004 | 1.613 | 1.896 | 0.251 | 0.090 | -0.104 | -0.151 |
| #24 | 0.075 | 0.152 | 0.815 | 1.621 | 0.002 | 0.004 | -1.043 | -0.091 | -0.924 | 0.276 | 0.840 | 0.592 |

Supplementary Table S3: **Subject-level intensity quality control for diffusion-derived microstructural metrics.** Subject-wise spatial mean and standard deviation of $\kappa$, $f_c$, and $\lambda_s$, computed across all voxels within the whole-brain diffusion mask, with corresponding $z$-scores.

| Outcome | $R^2$ | Adj. $R^2$ | $F$ | $p$ | $\beta_\kappa$ | $\beta_{f_c}$ | $SE_\kappa$ | $SE_{f_c}$ | $p_\kappa$ | $p_{f_c}$ |
|---|---|---|---|---|---|---|---|---|---|---|
| Complement | 0.869 | 0.816 | 16.512 | 0.006 | 0.819 | 0.204 | 0.183 | 0.183 | 0.007 | 0.316 |
| TREM2 | 0.869 | 0.816 | 16.552 | 0.006 | 0.863 | 0.131 | 0.183 | 0.183 | 0.005 | 0.505 |
| SALL1 | 0.704 | 0.585 | 5.939 | 0.048 | -0.015 | -0.832 | 0.275 | 0.275 | 0.959 | 0.029 |
| TGF | 0.650 | 0.511 | 4.650 | 0.072 | 0.159 | -0.868 | 0.299 | 0.299 | 0.618 | 0.034 |
| P2RY12 | 0.319 | 0.046 | 1.170 | 0.383 | -0.238 | -0.412 | 0.417 | 0.417 | 0.593 | 0.368 |
| TMEM119 | 0.464 | 0.249 | 2.163 | 0.211 | -0.371 | -0.423 | 0.370 | 0.370 | 0.362 | 0.305 |
| APOE | 0.254 | -0.044 | 0.851 | 0.481 | 0.278 | 0.310 | 0.437 | 0.437 | 0.552 | 0.510 |
| AQP4 | 0.175 | -0.155 | 0.531 | 0.618 | -0.097 | -0.364 | 0.459 | 0.459 | 0.841 | 0.464 |

| Outcome | $CI_{\kappa,L}$ | $CI_{\kappa,U}$ | $CI_{f_c,L}$ | $CI_{f_c,U}$ | Boot $CI_{\kappa,L}$ | Boot $CI_{\kappa,U}$ | Boot $CI_{f_c,L}$ | Boot $CI_{f_c,U}$ | $p_{boot,\kappa}$ | $p_{boot,f_c}$ | Partial $R^2_\kappa$ | Partial $R^2_{f_c}$ | $r_{obs,pred}$ | $p_{obs,pred}$ |
|---|---|---|---|---|---|---|---|---|---|---|---|---|---|---|
| Complement | 0.347 | 1.290 | -0.267 | 0.676 | 0.477 | 1.448 | -0.392 | 0.597 | 0.006 | 0.310 | 0.799 | 0.199 | 0.932 | 0.001 |
| TREM2 | 0.392 | 1.334 | -0.340 | 0.602 | -0.026 | 1.229 | -0.399 | 0.464 | 0.054 | 0.516 | 0.816 | 0.093 | 0.932 | 0.001 |
| SALL1 | -0.723 | 0.693 | -1.539 | -0.124 | -0.408 | 1.058 | -1.480 | 0.170 | 0.974 | 0.068 | 0.001 | 0.646 | 0.839 | 0.009 |
| TGF | -0.610 | 0.928 | -1.637 | -0.100 | -0.546 | 0.907 | -1.479 | -0.271 | 0.608 | 0.023 | 0.053 | 0.628 | 0.806 | 0.016 |
| P2RY12 | -1.311 | 0.835 | -1.486 | 0.661 | -0.945 | 1.124 | -2.616 | 0.416 | 0.759 | 0.344 | 0.061 | 0.163 | 0.565 | 0.145 |
| TMEM119 | -1.323 | 0.581 | -1.375 | 0.529 | -1.168 | 0.656 | -3.441 | 0.069 | 0.547 | 0.084 | 0.167 | 0.207 | 0.681 | 0.063 |
| APOE | -0.845 | 1.401 | -0.813 | 1.433 | -0.365 | 1.607 | -1.069 | 2.031 | 0.318 | 0.600 | 0.075 | 0.092 | 0.504 | 0.203 |
| AQP4 | -1.278 | 1.084 | -1.545 | 0.816 | -1.304 | 1.056 | -1.868 | 0.620 | 0.774 | 0.469 | 0.009 | 0.112 | 0.419 | 0.302 |

**Supplementary Table S4**

**Multiple regression statistics relating diffusion-derived microstructural metrics to HBCA transcriptional signatures across eight macro-regions.** Associations between $\kappa$ and $f_c$ and HBCA-derived transcriptional measures were assessed using multiple linear regression across eight macro-regions (n = 8). The table reports full model statistics (F-test, $p$-value, $R^2$, adjusted $R^2$), standardised regression coefficients ($\beta$), standard errors, 95% confidence intervals, partial $R^2$ values, observed–predicted correlation ($r$), and bootstrap-derived p-values and confidence intervals (10,000 iterations). Models are shown for complement-associated module, TREM2, SALL1, APOE, AQP4, and additional markers (TGF, P2RY12, TMEM119). For all models, numerator degrees of freedom were $df = 2$ (two predictors) and denominator degrees of freedom were $df = 5(n - k - 1)$, where $n = 8$ regions and $k = 2$ predictors).

| Metric | Region | Male Mean | Female Mean | Direction | $F(1,20)$ | $p$ | $p_{\mathrm{FDR}}$ |
|---|---|---|---|---|---|---|---|
| $\kappa$ | Cerebral cortex | 2.518 | 2.541 | Female > Male | 0.145 | 0.707 | 0.943 |
| | Cerebellum | 2.532 | 2.565 | Female > Male | 0.493 | 0.491 | 0.785 |
| | Thalamus | 3.189 | 3.090 | Male > Female | 1.152 | 0.296 | 0.740 |
| | Basal ganglia | 2.802 | 2.810 | Female > Male | 0.005 | 0.944 | 0.944 |
| | Hippocampal formation | 2.647 | 2.650 | Female > Male | 0.005 | 0.944 | 0.944 |
| | Amygdala | 2.455 | 2.514 | Female > Male | 0.842 | 0.370 | 0.740 |
| | Midbrain | 3.707 | 3.565 | Male > Female | 1.894 | 0.184 | 0.736 |
| | Pons and medulla | 4.125 | 3.854 | Male > Female | 6.792 | 0.017 | 0.135 |
| $f_c$ | Cerebral cortex | 0.270 | 0.267 | Male > Female | 0.242 | 0.628 | 0.838 |
| | Cerebellum | 0.325 | 0.321 | Male > Female | 0.080 | 0.780 | 0.891 |
| | Thalamus | 0.320 | 0.334 | Female > Male | 3.393 | 0.080 | 0.214 |
| | Basal ganglia | 0.339 | 0.347 | Female > Male | 3.587 | 0.073 | 0.214 |
| | Hippocampal formation | 0.281 | 0.288 | Female > Male | 1.525 | 0.231 | 0.463 |
| | Amygdala | 0.317 | 0.318 | Female > Male | 0.013 | 0.910 | 0.910 |
| | Midbrain | 0.313 | 0.322 | Female > Male | 0.988 | 0.332 | 0.531 |
| | Pons and medulla | 0.325 | 0.350 | Female > Male | 4.374 | 0.049 | 0.214 |
| Complement Module | Cerebral cortex | 2.117 | 2.135 | Female > Male | 0.141 | 0.712 | 0.929 |
| | Cerebellum | 2.140 | 2.166 | Female > Male | 0.472 | 0.500 | 0.800 |
| | Thalamus | 2.677 | 2.598 | Male > Female | 1.098 | 0.307 | 0.748 |
| | Basal ganglia | 2.364 | 2.372 | Female > Male | 0.008 | 0.929 | 0.929 |
| | Hippocampal formation | 2.225 | 2.229 | Female > Male | 0.012 | 0.915 | 0.929 |
| | Amygdala | 2.075 | 2.123 | Female > Male | 0.826 | 0.374 | 0.748 |
| | Midbrain | 3.099 | 2.985 | Male > Female | 1.835 | 0.191 | 0.748 |
| | Pons and medulla | 3.444 | 3.227 | Male > Female | 6.287 | 0.021 | 0.167 |
| TREM2 Map | Cerebral cortex | 2.210 | 2.229 | Female > Male | 0.142 | 0.710 | 0.935 |
| | Cerebellum | 2.229 | 2.257 | Female > Male | 0.480 | 0.496 | 0.794 |
| | Thalamus | 2.796 | 2.712 | Male > Female | 1.119 | 0.303 | 0.745 |
| | Basal ganglia | 2.464 | 2.472 | Female > Male | 0.007 | 0.935 | 0.935 |
| | Hippocampal formation | 2.322 | 2.326 | Female > Male | 0.009 | 0.926 | 0.935 |
| | Amygdala | 2.162 | 2.213 | Female > Male | 0.832 | 0.372 | 0.745 |
| | Midbrain | 3.242 | 3.121 | Male > Female | 1.858 | 0.188 | 0.745 |
| | Pons and medulla | 3.605 | 3.374 | Male > Female | 6.482 | 0.019 | 0.154 |
| SALL1 Map | Cerebral cortex | -0.263 | -0.260 | Female > Male | 0.233 | 0.635 | 0.811 |
| | Cerebellum | -0.308 | -0.305 | Female > Male | 0.059 | 0.811 | 0.811 |
| | Thalamus | -0.314 | -0.324 | Male > Female | 3.310 | 0.084 | 0.335 |
| | Basal ganglia | -0.324 | -0.331 | Male > Female | 3.444 | 0.078 | 0.335 |
| | Hippocampal formation | -0.273 | -0.279 | Male > Female | 1.609 | 0.219 | 0.439 |
| | Amygdala | -0.301 | -0.302 | Male > Female | 0.072 | 0.791 | 0.811 |
| | Midbrain | -0.316 | -0.321 | Male > Female | 0.434 | 0.518 | 0.811 |
| | Pons and medulla | -0.332 | -0.349 | Male > Female | 2.519 | 0.128 | 0.342 |

**Supplementary Table S5**
**Sex-related differences across diffusion-derived microstructural metrics and regression-weighted maps.** Sex-related effects were evaluated at the macro-regional level (n = 8 regions) using ANCOVA controlling for age. The table reports male and female means, direction of effect, F-statistics, uncorrected p-values, and false discovery rate (FDR)-adjusted p-values for $\kappa$, $f_c$, the complement-associated module–derived map, TREM2-derived map, and SALL1-derived map. FDR correction was applied across the eight macro-regions within each metric. No comparisons survived FDR correction ($q < 0.05$).

#### C. Supplemental Figures

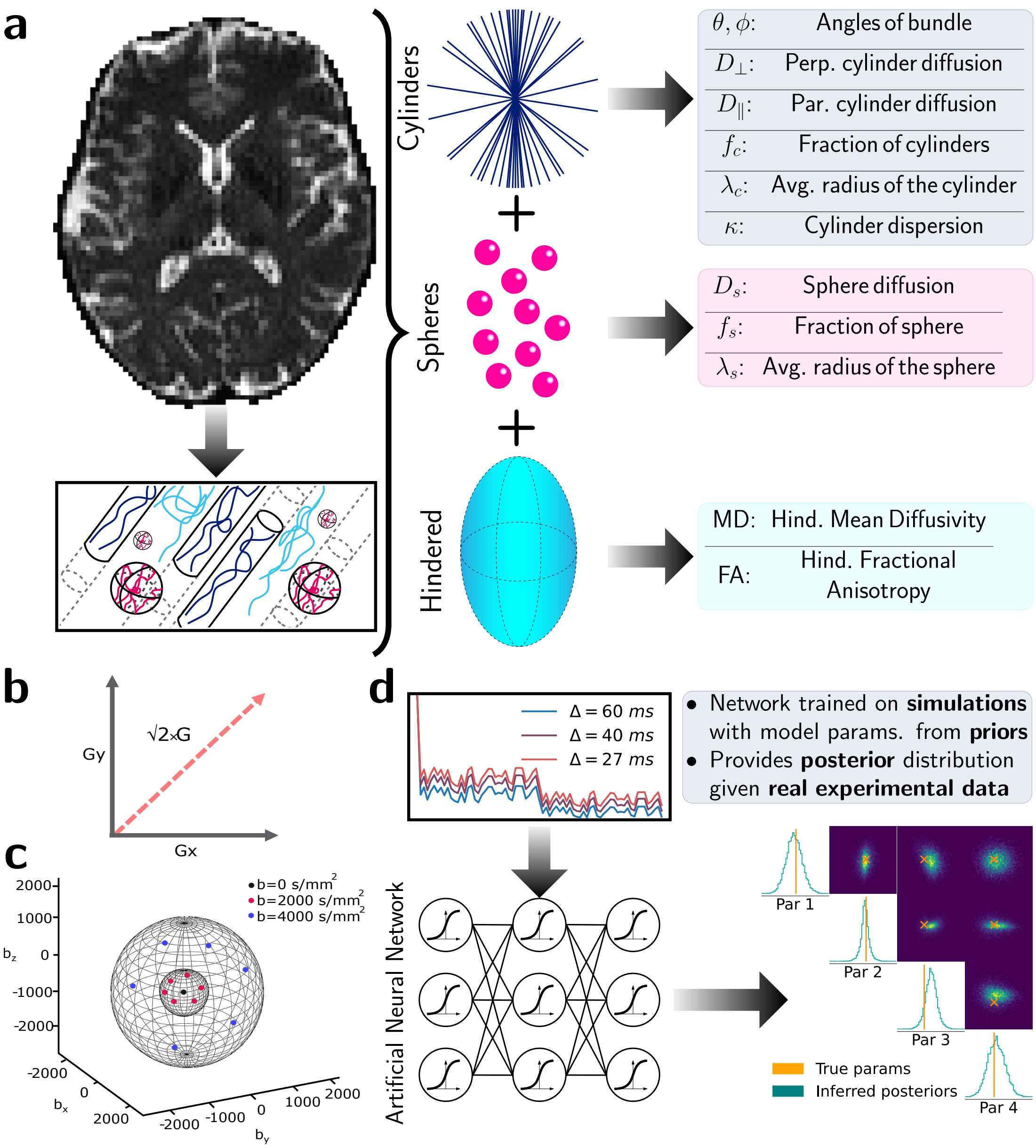


**Supplementary Figure S1: Multi–diffusion-time modelling framework for estimating microstructural parameters in the human brain using advanced acquisition and simulation-based inference techniques. a)** Schematic representation of the stick-and-ball microstructural model comprising three diffusion environments: (i) a hindered compartment, (ii) a spherical compartment representing soma-like structures; and (iii) a cylindrical compartment representing radially symmetric processes. **b)** Orthogonal gradient composition used to increase achievable diffusion weighting. **c)** Angular sampling scheme of the gradient orientation used for diffusion encoding. **d)** Sampling across gradient strengths, gradient directions, and diffusion times produces diffusion signals that encode a characteristic fingerprint of the underlying microstructure. These signals are provided to a neural network pre-trained on simulated data generated by the multi-compartment model, which outputs the posterior distribution over model parameters conditioned on the measured signal.

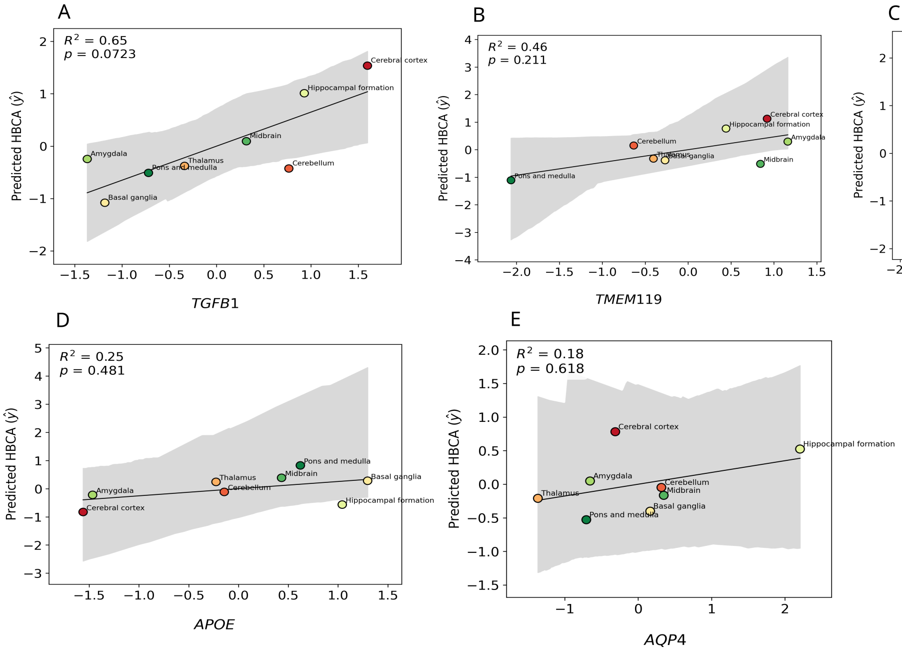


**Supplementary Figure S2:** Multiple linear regression models across eight macro-regions (n = 8) relating $\kappa$ and $f_c$ to HBCA-derived transcriptional measures for **a)** TGF, **b)** TMEM119, **c)** P2RY12, **d)** APOE (astrocyte-associated control marker), and **e)** AQP4 (astrocyte-associated control marker). Each point represents one macro-region. Solid black lines indicate the fitted regression relationship, and shaded regions represent 95% bootstrapped confidence intervals. Model statistics ($R^2$ and $F$-test $p$-values) are displayed in each panel.

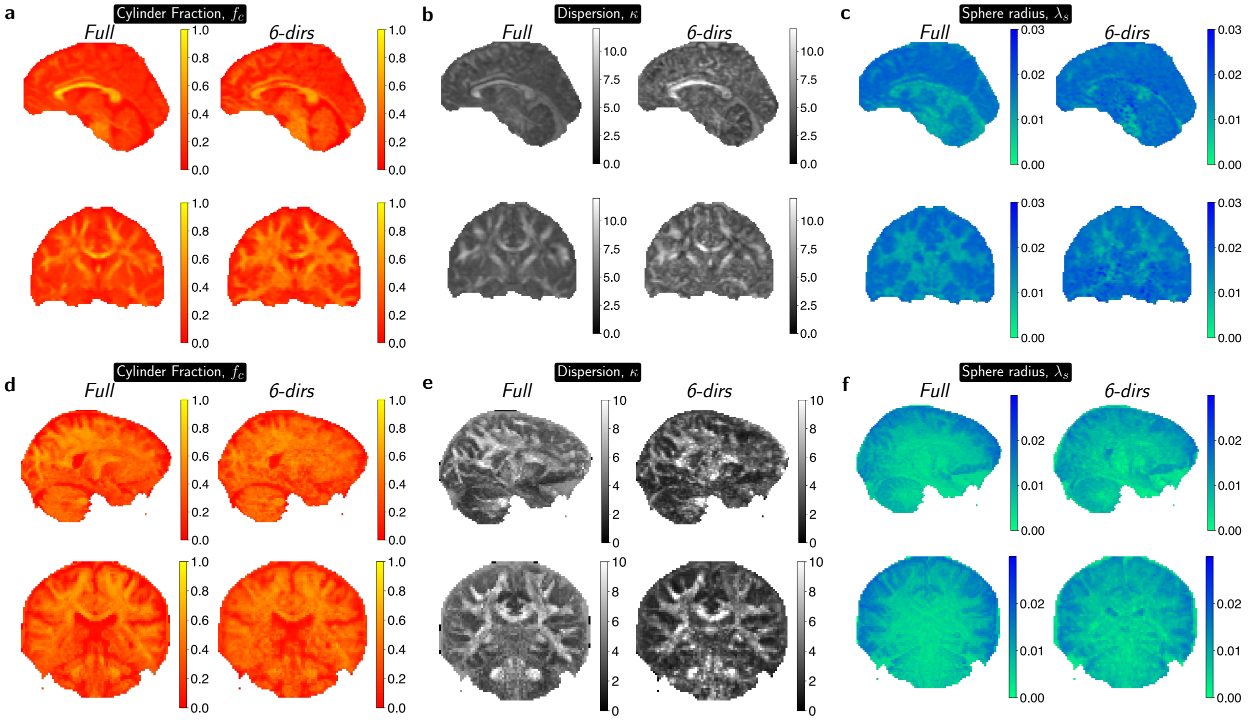


**Supplementary Figure S3: Alternative planes of slices shown in Fig. 2. a-c)** Sagittal and coronal slices of the same subject of Fig. 2a-b from the Garcia-Hernandez et al. (2022) dataset shown for completeness. a,b and c show cylinder fraction, dispersion and sphere radius, respectively. **d-f)** Similar to a-c, but for our newly generated dataset in Figs. 2a-b.

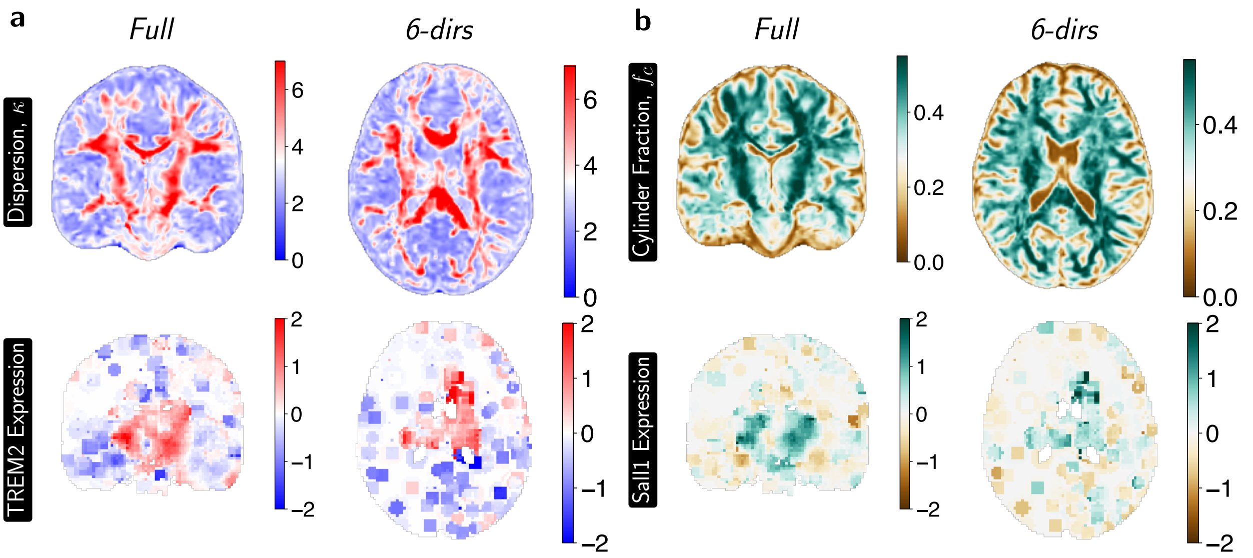


**Supplementary Figure S4: Alternative planes of slices shown in Fig. 4. a)** Coronal and axial slices of the cohort-averaged $\kappa$ and TREM2 expression as shown of Fig. 4g. **b)** Similar to a, but for cohort-averaged $f_c$ and Sall1 expression
